# Investigating the causal interplay between sleep traits and risk of acute myocardial infarction: a Mendelian randomization study

**DOI:** 10.1101/2023.04.17.23288680

**Authors:** Nikhil Arora, Laxmi Bhatta, Eivind Schjelderup Skarpsno, Håvard Dalen, Bjørn Olav Åsvold, Ben Michael Brumpton, Rebecca Claire Richmond, Linn Beate Strand

## Abstract

**Background:** Few studies have investigated the joint effects of sleep traits on the risk of acute myocardial infarction (AMI). No previous study has used factorial Mendelian randomization (MR) which may reduce confounding, reverse causation and measurement error. Thus, it is prudent to study joint effects using robust methods to propose sleep-targeted interventions which lower the risk of AMI.

**Methods:** The causal interplay between combinations of two sleep traits (including insomnia symptoms, sleep duration or chronotype) on the risk of AMI was investigated using factorial MR. Genetic risk scores for each sleep trait were dichotomized at their median in UK Biobank (UKBB) and the second survey of the Trøndelag Health Study (HUNT2). A combination of two sleep traits constituting 4 groups were analysed to estimate the risk of AMI in each group using a 2×2 factorial MR design.

**Results:** In UKBB, participants with high genetic risk for both insomnia symptoms and short sleep had the highest risk of AMI (hazard ratio (HR) 1.10; 95% confidence interval (CI) 1.03, 1.18), although there was no evidence of interaction (relative excess risk due to interaction (RERI) 0.03; 95% CI −0.07, 0.12). These estimates were less precise in HUNT2 (HR 1.02; 95% CI 0.93, 1.13), possibly due to weak instruments. Participants with high genetic risk for both a morning chronotype and insomnia symptoms (HR 1.09; 95% CI 1.03, 1.17); and a morning chronotype and short sleep (HR 1.11; 95% CI 1.04, 1.19) had the highest risk of AMI in UKBB, although there was no evidence of interaction (RERI 0.03; 95% CI −0.06, 0.12; and RERI 0.05; 95% CI −0.05, 0.14, respectively). Chronotype was not available in HUNT2.

**Conclusions:** This study reveals no interaction effects between sleep traits on the risk of AMI, but all combinations of sleep traits increased the risk of AMI except those with long sleep. This indicates that the main effects of sleep traits on AMI are likely to be independent of each other.

## Background

Poor sleep is a major public health problem that has emerged as being associated with several health conditions [1, 2], including those related to cardiovascular health such as hypertension [2, 3], obesity [2, 4], and dyslipidaemia [5]. Cardiovascular diseases (CVDs) account for a large part of global morbidity, and are the leading cause of death [6]. Since sleep problems can be managed through cognitive-behavioural therapy and medication [7], understanding how sleep impacts cardiovascular health can have important implications for interventions that aim to target sleep with an objective to lower the risk of CVDs.

Sleep is a complex and multifaceted biological phenomenon which comprises several traits [8]. Previous observational studies have mainly focused on individual sleep traits as separate risk factors for CVDs [9–13]. Insomnia symptoms, short or long sleep duration, and evening chronotype have been identified as individual risk factors for acute myocardial infarction (AMI) [9, 11, 13, 14]. Sleep traits are often correlated and can together assert their influence on the disease risk. Few observational studies have investigated the joint effects of sleep traits and have found evidence that sleep traits interact to increase the risk of cardiovascular outcomes [14–20]. For instance, insomnia with short sleep considered the most biologically severe sleep disorder phenotype [21], has been found to be associated with increased cardiometabolic risk [14, 16, 18–20]. In our recent study, we observed that those reporting two sleep traits (including insomnia symptoms, short sleep, long sleep and evening chronotype) had a higher incidence of AMI than those reporting only one sleep trait. Any relative excess risk due to interaction (RERI) was only observed among those reporting insomnia symptoms and long sleep duration [14]. However, the available evidence on the joint effects of sleep traits on the risk of AMI is based on conventional observational studies that are prone to bias due to residual confounding, reverse causation and measurement error [22].

Mendelian randomization (MR) uses genetic variants as instruments that are robustly associated with a modifiable risk factor to investigate the causal effect on an outcome [23]. MR exploits the fact that genetic variants are randomly assigned to individuals and fixed at conception, making it less susceptible to the bias observed in conventional observational studies. Recent MR studies have evaluated individual effects of sleep traits on CVDs, providing evidence of an adverse effect of insomnia symptoms on prevalent coronary artery disease (CAD) [24–27] and AMI [28], and a protective effect per hour increase in sleep duration and an adverse effect of short sleep on CAD and AMI [11, 29] (see a summary in Additional file 1: Table S1) [11, 24–33]. MR investigation of chronotype is scarce and lacks compelling evidence [28], thus it remains unclear whether chronotype itself is causally associated with an increased risk of AMI, or if the adverse effect of circadian preference can be explained by insomnia symptoms or sleep duration. More importantly, MR investigations exploring the joint causal effects of sleep traits on risk of AMI remain largely untapped, which could provide robust evidence on the risk of AMI from experiencing two sleep traits simultaneously.

In this study, we therefore used one-sample and factorial MR to investigate the causal effects of individual sleep traits (insomnia symptoms, sleep duration and chronotype) and their joint effects on incident AMI, in two large longitudinal studies (UK Biobank (UKBB) and the second survey of the Trøndelag Health Study (HUNT2)).

## Methods

### Study participants

#### UK Biobank

Out of 9.2 million eligible adults (ranging between 40 and 70 years) in the UK who were invited to participate, more than 500 000 participated in the study during March 2006 – July 2010 (5.5% response rate). The participants visited one of the 22 study assessment centres located throughout England, Scotland and Wales, where they signed an electronic consent and completed a touchscreen questionnaire along with a brief computer-assisted interview. They provided detailed information about their lifestyle, physical measures and had blood, urine and saliva samples collected and stored for future analysis, as described elsewhere [34]. The UKBB received approval from the National Health Service (NHS) Research Ethics Service (reference number 11/NW/0382) and the database was created in compliance with the Declaration of Helsinki.

#### HUNT study

All inhabitants aged 20 years or older in the Nord-Trøndelag region of Norway, were invited to participate in a four-phase population-based health survey (the HUNT study), first in 1984-86 (HUNT1), then in 1995-97 (HUNT2) and 2006-08 (HUNT3), and last in 2017-19 (HUNT4). This study is based on data from HUNT2, where 93 898 individuals were invited and 65 228 (69.5%) participated [35]. The invitation letter was sent by mail along with a self-administered questionnaire. The participants attended examination stations where clinical examination was performed, and blood samples were drawn by trained personnel. Detailed information regarding HUNT2 study has been published elsewhere [36]. The HUNT Study was approved by the Data Inspectorate of Norway and recommended by the Regional Committee for Ethics in Medical Research (REK; reference number 152/95/AH/JGE). Additionally, the ethical clearance for conducting this study was obtained from the Regional Committee for Ethics in Medical Research (REK nord; reference number 2020/47206).

### Sleep traits

#### Insomnia symptoms

In both UKBB and HUNT2, insomnia symptoms were defined as two night-time insomnia symptoms (i.e., difficulty falling asleep, difficulty maintaining sleep or waking up too early) without information about daytime impairment. Thus our definition for insomnia symptoms did not include all components used in the frameworks for diagnosing insomnia [37].

In UKBB, participants were asked: “Do you have trouble falling asleep at night or do you wake up in the middle of the night?” with response options “Never/rarely”, “Sometimes”, “Usually” or “Prefer not to answer”. Participants were classified as having insomnia symptoms if they answered “Usually”; and not having insomnia symptoms if they answered “Never/rarely” or “Sometimes”. Other responses were coded as missing.

In HUNT2, insomnia symptoms were assessed by the following two questions: “Have you had difficulty falling asleep in the last month?”, and “During the last month, have you woken too early and not been able to get back to sleep?” with response options “Never”, “Sometimes”, “Often” or “Almost every night”. Participants who responded “Often” or “Almost every night” to at least one of these questions were classified as having insomnia symptoms. For participants who answered only one of these insomnia symptom questions, we did the following: (1) if they answered “Often” or “Almost every night” to one of the questions, but did not answer the other, they were classified as having insomnia symptoms, and (2) if they answered “Never” or “Sometimes” to one of the questions, but did not answer the other, they were excluded to avoid possible misclassification. The remaining participants were classified as not having insomnia symptoms.

#### Sleep duration

Sleep duration was assessed by the questions: “About how many hours sleep do you get in every 24 hours? (please include naps)” and “How many hours do you usually spend lying down (i.e. sleeping and/or napping) during a 24-hour period?” in UKBB and HUNT2, respectively. The answers could only contain integer values. Any influence of poor health on implausible short or long sleep durations was avoided by excluding extreme responses of less than 3 hours or more than 18 hours. Binary variables for short sleep (≤6 hours vs. 7-8 hours) and long sleep (≥9 hours vs. 7-8 hours) were also constructed.

#### Chronotype

Chronotype (morning or evening chronotype) in UKBB was assessed by the question: “Do you consider yourself to be?” with response options “Definitely a ‘morning’ person”, “More a ‘morning’ than ‘evening’ person”, “More an ‘evening’ than a ‘morning’ person”, “Definitely an ‘evening’ person”, “Do not know”, or “Prefer not to answer”. Participants were classified as having a morning chronotype if they reported “Definitely a ‘morning’ person” or “More a ‘morning’ than ‘evening’ person”, and as having an evening chronotype if they reported “More an ‘evening’ than a ‘morning’ person” or “Definitely an ‘evening’ person”. Other responses were coded as missing. Chronotype was not reported in any survey of the HUNT Study.

### Acute Myocardial Infarction (AMI)

In UKBB, participants were followed through record linkage to the Hospital Episode Statistics (HES) for England, Scottish Morbidity Record (SMR) and Patient Episode Database for Wales (PEDW) where health-related outcomes had been defined by International Classification of Diseases (ICD)-9 and ICD-10 codes (Field IDs: 41270, 41271, 41280 and 41281). Also, mortality records were obtained from the NHS Digital for participants in England and Wales, and from the NHS Central Register (part of the National Records of Scotland) for participants in Scotland where cause of death had been defined by ICD-10 codes (Field IDs: 40001 and 40000).

In HUNT2, participants were followed via linkage to the medical records from the three hospitals (St. Olavs Hospital, Levanger Hospital and Namsos Hospital) of the Nord-Trøndelag region where health-related outcomes had been defined by ICD-9 and ICD-10 codes. Mortality records were identified by a linkage to the National Cause of Death Registry where cause of death had been defined by ICD-10 codes.

Any hospitalization or death due to AMI were identified using ICD-9 code 410, and ICD-10 codes I21 and I22. Each participant was followed until either first diagnosis/death due to AMI, death due to other cause, loss to follow-up, or end of follow-up (March 23, 2021 for UKBB and Dec. 31, 2020 for HUNT2). Incident cases were defined as the first occurrence of either hospitalization or death due to AMI during follow-up. Participants with any previous AMI episode(s) before their date of participation in the study regarded as prevalent cases, were excluded in the study.

### Covariates

Several factors to be potential confounders of the exposure-outcome relation were considered. The covariates selected a priori were age, gender, marital status (married, unmarried or separated/divorced/widowed), frequency of alcohol intake (never, monthly, weekly, or daily), smoking history (never, ex-smoker or current smoker), body mass index (BMI), level of physical activity (inactive/low, moderate, or high), Townsend deprivation index (TDI; for UKBB only), education attainment (≤10 years, 11-13 years, or ≥14 years), shift work (yes or no), employment status (employed or not employed), systolic blood pressure (SBP), blood cholesterol levels, blood glucose levels, depression (yes or no in UKBB; and Hospital Anxiety and Depression Scale (HADS) – Depression scores in HUNT2), anxiety (yes or no in UKBB; and HADS – Anxiety scores in HUNT2), use of sleep medication (yes or no) and chronic illness (yes or no). The details on how covariates were handled are described in the supplementary material (see Additional file 1) [36, 38–47].

### Genetic variants

In UKBB, participants were genotyped using either one of the UK BiLEVE or the UK Biobank Axiom genotyping chips. The genetic variants used were extracted genotypes from the UK Biobank imputation dataset (imputed to the UK10K plus 1000 Genomes phase 3 and Haplotype Reference Consortium reference panels), that were quality controlled using a standard protocol [48, 49]. In HUNT, participants were genotyped with one of three different Illumina HumanCoreExome genotyping chips (HumanCoreExome 12 v.1.0, HumanCoreExome 12 v.1.1, and UM HUNT Biobank v.1.0), where genotypes from different chips were quality controlled separately and reduced to a common set of variants. The quality control measures used were similar to UKBB [50]. All genotyped samples included were of European decent.

A total of 248 single nucleotide polymorphisms (SNPs) were identified as robustly associated with insomnia symptoms [30], 78 SNPs associated with 24-hour sleep duration [31], and 351 SNPs associated with morning preference chronotype [32], at a genome-wide significance level (P <5×10^-8^) from three large genome-wide association studies (GWASs). In addition, 27 and 8 SNPs were identified to associate with short and long sleep duration, respectively [31]. The detailed information about discovery GWASs from where genetic instruments were identified were listed in Table 1.

**Table 1:** Summary of genome-wide significant genetic instruments of sleep traits in the discovery genome-wide association studies.

| Sleep traits | Discovery GWAS | PMID | N | Cohorts used by the discovery GWAS |  | No. of SNPs identified |
| --- | --- | --- | --- | --- | --- | --- |
|  |  |  |  | UKBB | 23andMe |  |
| Insomnia symptoms | Jansen *et al.*, 2019 [30] | 30804565 | 1 331 010 | 109 402 cases and 277 131 controls | 288 557 cases and 655 920 controls | 248 |
| 24-hour sleep duration (h) | Dashti *et al.*, 2019 [31] | 30846698 | 446 118 | 446 118 samples | Not included | 78 |
| Short sleep ($\leq 6$ h vs. 7-8 h) | Dashti *et al.*, 2019 [31] | 30846698 | 411 934 | 106 192 cases and 305 742 controls | Not included | 27 |
| Long sleep ($\geq 9$ h vs. 7-8 h) | Dashti *et al.*, 2019 [31] | 30846698 | 339 926 | 34 184 cases and 305 742 controls | Not included | 8 |
| Chronotype (morning preference)* | Jones *et al.*, 2019 [32] | 30696823 | 651 295 | 252 287 cases and 150 908 controls | 120 478 cases and 127 622 controls | 351 |

GWAS indicated genome-wide association studies; N, sample size; SNPs, single nucleotide polymorphisms; and UKBB, UK Biobank.
\* In the discovery GWAS of chronotype, the chronotype increasing allele is morning preference.

### Statistical analysis

Genetic risk score (GRS) for each sleep trait were created as an instrument that could overcome the weak effect of most SNPs on their corresponding sleep trait [51]. Weighted GRS (wGRS) were calculated as the sum of the participants’ sleep trait increasing alleles (morning preference alleles for chronotype; thus evening chronotype as reference), weighted by the variant effect sizes from the external GWAS. wGRS were incorporated for our main analysis in HUNT2 only, whereas in UKBB, we used unweighted GRS (uwGRS) calculated as sum of the sleep trait increasing alleles. Since all included discovery GWASs used the UKBB cohort, the use of internal weights to calculate wGRS is not recommended [51].

Instrument strength was assessed by regressing each sleep trait on their respective GRS and reporting R^2^ and F-statistics. The causal effects of individual sleep traits (insomnia symptoms, 24-hour sleep duration, short sleep, long sleep and chronotype) on the risk of incident AMI were tested using a one-sample MR analysis. A factorial MR analysis was used to investigate the joint causal effects of any two sleep traits (i.e., insomnia symptoms and short sleep, or insomnia symptoms and long sleep, or insomnia symptoms and chronotype, or short sleep and chronotype, or long sleep and chronotype) on the risk of incident AMI. All analyses were conducted using R version 3.6.3 (R Foundation for Statistical Computing, Vienna, Austria).

#### One-sample MR analysis

One-sample MR analysis was performed for each sleep trait using individual-level data separately in UKBB and HUNT2. A two-stage predictor substitution (TSPS) regression estimator method was used to calculate average causal hazard ratios (HRs). The first stage involved regression of each sleep trait (linear regression for 24-hour sleep duration, and logistic regression for other sleep traits) on their GRS, and the second stage consisted of a Cox regression of AMI status on the fitted values from the first stage regression, with adjustment for age at recruitment, gender, assessment centre (in UKBB), genetic principal components (40 in UKBB and 20 in HUNT2), and genotyping chip in both stages. As recommended for MR analysis with a binary outcome [52], the first stage regression was restricted to participants who did not experience AMI. To obtain corrected standard errors, a bootstrapping method was applied with 2000 iterations in UKBB and 5000 iterations in HUNT2 [52]. The causal estimates for insomnia symptoms, short sleep, long sleep and chronotype were scaled to represent the risk increase in AMI per doubling in the odds of these exposures, by multiplying the obtained β values by 0.693 as previously described [53]. The causal estimate for 24-hour sleep duration represents the risk increase in AMI per additional hour of sleep.

#### Factorial MR analysis

A 2×2 factorial MR was applied where each of the sleep traits (except 24-hour sleep duration) was dichotomized at their median GRS (uwGRS for UKBB and wGRS for HUNT2), with values equal to or below the median represented low genetic risk for the sleep trait, and values above the median represented high genetic risk for the sleep trait. Thus, for any combination of two sleep traits, participants were categorized into 4 groups according to their genetic predisposition. For instance, when combining insomnia symptoms and short sleep, participants were categorized into: *“Both GRS ≤ median”* (reference; representing low genetic risk for both insomnia symptoms and short sleep), *“Insomnia GRS > median”* (representing high genetic risk for insomnia symptoms only), *“Short sleep GRS > median*” (representing high genetic risk for short sleep only) and *“Both GRS > median”* (representing high genetic risk for both insomnia symptoms and short sleep). Cox regression was then used to investigate the association between these groups and incident AMI, with adjustment for age at recruitment, gender, assessment centre (in UKBB), genetic principal components (40 in UKBB and 20 in HUNT2), and genotyping chip. Furthermore, interaction between any two sleep traits on risk of AMI was assessed by calculating relative excess risk due to interaction (RERI) using the risk estimates obtained for each sleep trait combination when none of the HRs were less than 1 (i.e. preventive) [54, 55]. RERI equals 0 implies exact additivity (no interaction), RERI >0 implies more than additivity (positive interaction or synergism), and RERI <0 implies less than additivity (negative interaction or antagonism).

#### Sensitivity analyses

To check the proportionality of hazards, the Pearson’s correlations were used to test Schoenfeld residuals from one-sample MR and 2×2 factorial MR Cox regression models for an association with follow-up time. To check the robustness of the findings, the one-sample MR and 2×2 factorial MR analyses were repeated using uwGRS in HUNT2.

To assess the second MR assumption that the genetic instruments used are independent of confounders, associations of the GRS and potential confounders were investigated in UKBB and HUNT2. Furthermore, one-sample MR analysis adjusted for any potential confounders found strongly associated with the sleep trait GRS in two cohorts (beyond a Bonferroni significance threshold of P <5.88×10^-4^ in UKBB and P <7.81×10^-4^ in HUNT2) were performed.

To investigate potential directional pleiotropy, the estimates of the SNP-exposure and SNP-outcome associations from the same participants were obtained and two-sample MR methods, such as MR-Egger, weighted median and weighted mode-based methods were applied. Each of these methods makes different assumptions about the genetic instruments used, where the MR-Egger regression method gives a valid causal estimate under the InSIDE (instrument strength independent of direct effect) assumption and its intercept allows the size of any unbalanced pleiotropic effect to be determined [56], weighted median method assumes at least 50% of genetic variants are valid [57], and weighted mode-based estimation method assumes a plurality of genetic variants are valid [58]. These methods can be applied in a one-sample setting [59], and consistent estimates across these methods strengthens causal evidence. To further investigate pleiotropy due to insomnia symptoms’ instruments, 57 SNPs found robustly associated with insomnia symptoms by Lane *et al.* [24] in another GWAS on UKBB (n = 345 022 cases and 108 357 controls) representing crucial variants with effect sizes for any insomnia symptoms (“sometimes”/“usually” as cases versus “never/rarely” as controls), were used in a post hoc one-sample MR Cox regression analysis using different methods.

To evaluate potential impact of winner’s curse, one-sample MR analysis was repeated using genetic variants that replicated at a genome-wide significance level (P <5×10^-8^) in a large independent dataset for insomnia symptoms (23andMe, n = 944 477; see Additional file 2: Table G1) [30] and chronotype (23andMe, n = 240 098; see Additional file 2: Table G5) [32].

As an additional analysis, continuous factorial MR analysis using two GRS (for any combination of two sleep traits) as quantitative traits and their product term was applied, to avoid potential bias due to arbitrary dichotomization and to maximize power [60]. Further, RERI was calculated as test of interaction using the risk estimates for the quantitative GRS and their product term for each sleep trait combination when none of the HRs were less than 1 (i.e. preventive) for AMI [54, 61].

As use of sleep medication has been associated with CVDs [62], one-sample MR (without applying bootstrap method) and 2×2 factorial MR analyses were repeated excluding participants who reported use of sleep medication(s).

## Results

Among the 336 262 participants in UKBB who passed the genetic quality control and had information available on the sleep traits, 11 399 (3.4%) had ever received the diagnosis of AMI. Of these, 3 586 (1.1%) prevalent cases with AMI diagnosis were excluded, and 7 813 (2.3%) had their first AMI diagnosis during a mean (standard deviation (SD)) follow-up of 11.7 (1.9) years (see Additional file 1: Figure S1). Among the 45 602 participants in HUNT2 who passed the genetic quality control and had information available for sleep traits of interest, 5 362 (11.7%) had ever received diagnosis of AMI. Of these, 874 (1.9%) prevalent cases with AMI diagnosis were excluded, and 4 488 (10.0%) had their first AMI diagnosis during a mean (SD) follow-up of 20.4 (6.9) years (see Additional file 1: Figure S1).

Table 2 represents the baseline characteristics of the study participants stratified by their AMI status in UKBB and HUNT2. Participants with an incidence of AMI during follow-up in the UKBB and HUNT2 were older and more likely to be males and current smokers. They were more likely to have used sleep medication(s), have a higher BMI, higher systolic blood pressure, higher blood glucose levels, and were suffering more from depression and chronic illness. They were also less likely to consume alcohol, be physically active, have a tertiary education and be employed compared to participants with no episodes of AMI. The HUNT2 participants with an AMI incidence during follow-up were more likely to have higher serum cholesterol levels, but less likely to be suffering from anxiety and working shifts in contrast to UKBB participants when compared to participants with no episode of AMI.

**Table 2:**
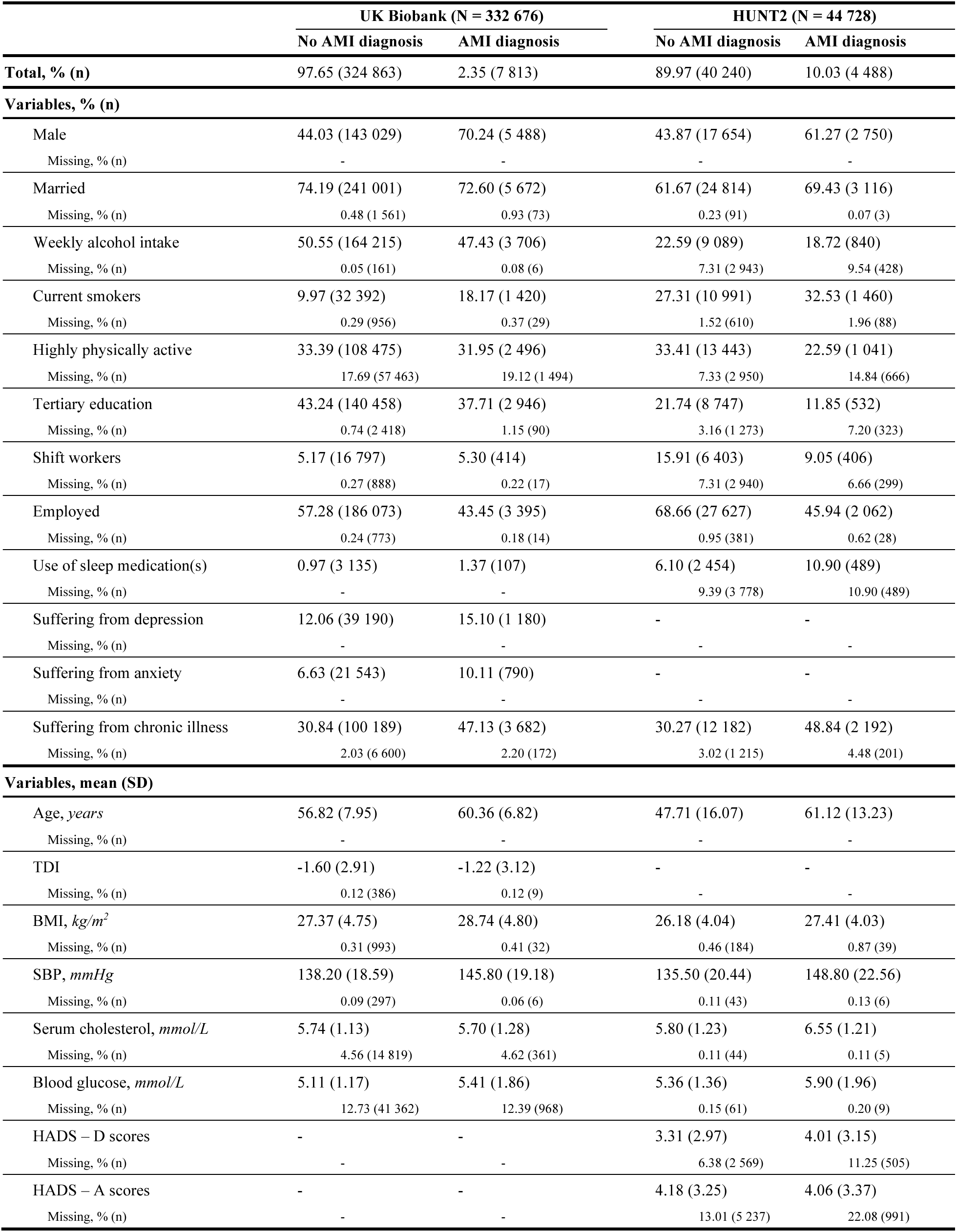
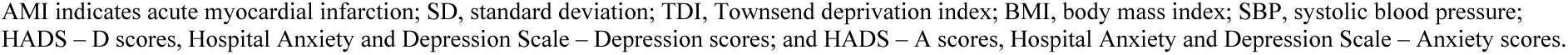
Baseline characteristics of study participants who had an episode of acute myocardial infarction (AMI) and not had AMI during follow-up in UK Biobank and HUNT2.

|  | UK Biobank (N = 332 676) |  | HUNT2 (N = 44 728) |  |
| --- | --- | --- | --- | --- |
|  | No AMI diagnosis | AMI diagnosis | No AMI diagnosis | AMI diagnosis |
| **Total, % (n)** | 97.65 (324 863) | 2.35 (7 813) | 89.97 (40 240) | 10.03 (4 488) |
| **Variables, % (n)** |  |  |  |  |
| Male | 44.03 (143 029) | 70.24 (5 488) | 43.87 (17 654) | 61.27 (2 750) |
| Missing, % (n) | - | - | - | - |
| Married | 74.19 (241 001) | 72.60 (5 672) | 61.67 (24 814) | 69.43 (3 116) |
| Missing, % (n) | 0.48 (1 561) | 0.93 (73) | 0.23 (91) | 0.07 (3) |
| Weekly alcohol intake | 50.55 (164 215) | 47.43 (3 706) | 22.59 (9 089) | 18.72 (840) |
| Missing, % (n) | 0.05 (161) | 0.08 (6) | 7.31 (2 943) | 9.54 (428) |
| Current smokers | 9.97 (32 392) | 18.17 (1 420) | 27.31 (10 991) | 32.53 (1 460) |
| Missing, % (n) | 0.29 (956) | 0.37 (29) | 1.52 (610) | 1.96 (88) |
| Highly physically active | 33.39 (108 475) | 31.95 (2 496) | 33.41 (13 443) | 22.59 (1 041) |
| Missing, % (n) | 17.69 (57 463) | 19.12 (1 494) | 7.33 (2 950) | 14.84 (666) |
| Tertiary education | 43.24 (140 458) | 37.71 (2 946) | 21.74 (8 747) | 11.85 (532) |
| Missing, % (n) | 0.74 (2 418) | 1.15 (90) | 3.16 (1 273) | 7.20 (323) |
| Shift workers | 5.17 (16 797) | 5.30 (414) | 15.91 (6 403) | 9.05 (406) |
| Missing, % (n) | 0.27 (888) | 0.22 (17) | 7.31 (2 940) | 6.66 (299) |
| Employed | 57.28 (186 073) | 43.45 (3 395) | 68.66 (27 627) | 45.94 (2 062) |
| Missing, % (n) | 0.24 (773) | 0.18 (14) | 0.95 (381) | 0.62 (28) |
| Use of sleep medication(s) | 0.97 (3 135) | 1.37 (107) | 6.10 (2 454) | 10.90 (489) |
| Missing, % (n) | - | - | 9.39 (3 778) | 10.90 (489) |
| Suffering from depression | 12.06 (39 190) | 15.10 (1 180) | - | - |
| Missing, % (n) | - | - | - | - |
| Suffering from anxiety | 6.63 (21 543) | 10.11 (790) | - | - |
| Missing, % (n) | - | - | - | - |
| Suffering from chronic illness | 30.84 (100 189) | 47.13 (3 682) | 30.27 (12 182) | 48.84 (2 192) |
| Missing, % (n) | 2.03 (6 600) | 2.20 (172) | 3.02 (1 215) | 4.48 (201) |
| **Variables, mean (SD)** |  |  |  |  |
| Age, years | 56.82 (7.95) | 60.36 (6.82) | 47.71 (16.07) | 61.12 (13.23) |
| Missing, % (n) | - | - | - | - |
| TDI | -1.60 (2.91) | -1.22 (3.12) | - | - |
| Missing, % (n) | 0.12 (386) | 0.12 (9) | - | - |
| BMI, kg/m<sup>2</sup> | 27.37 (4.75) | 28.74 (4.80) | 26.18 (4.04) | 27.41 (4.03) |
| Missing, % (n) | 0.31 (993) | 0.41 (32) | 0.46 (184) | 0.87 (39) |
| SBP, mmHg | 138.20 (18.59) | 145.80 (19.18) | 135.50 (20.44) | 148.80 (22.56) |
| Missing, % (n) | 0.09 (297) | 0.06 (6) | 0.11 (43) | 0.13 (6) |
| Serum cholesterol, mmol/L | 5.74 (1.13) | 5.70 (1.28) | 5.80 (1.23) | 6.55 (1.21) |
| Missing, % (n) | 4.56 (14 819) | 4.62 (361) | 0.11 (44) | 0.11 (5) |
| Blood glucose, mmol/L | 5.11 (1.17) | 5.41 (1.86) | 5.36 (1.36) | 5.90 (1.96) |
| Missing, % (n) | 12.73 (41 362) | 12.39 (968) | 0.15 (61) | 0.20 (9) |
| HADS – D scores | - | - | 3.31 (2.97) | 4.01 (3.15) |
| Missing, % (n) | - | - | 6.38 (2 569) | 11.25 (505) |
| HADS – A scores | - | - | 4.18 (3.25) | 4.06 (3.37) |
| Missing, % (n) | - | - | 13.01 (5 237) | 22.08 (991) |

Among UKBB participants, the variance explained by the uwGRS in insomnia symptoms, 24-hour sleep duration (h), short sleep (≤6 h vs. 7-8 h), long sleep (≥9 h vs. 7-8 h) and morning chronotype were 0.41%, 0.59%, 0.18%, 0.11% and 1.54%, respectively, and corresponding F-statistics were 1370.92, 1962.0, 558.68, 285.42 and 5202.20 (see Additional file 1: Table S2). The variance explained by the wGRS among HUNT2 participants in insomnia symptoms, 24-hour sleep duration, short sleep and long sleep were 0.16%, 0.09%, 0.01% and 0.01%, respectively, and the corresponding F-statistics were 71.17, 38.94, 4.97 and 4.07 (see Additional file 1: Table S2).

### Causal effects of individual sleep traits on the risk of AMI

There was evidence for an adverse causal effect on AMI risk per doubling in odds of insomnia symptoms in UKBB (HR 1.18; 95% CI 1.07, 1.31) and HUNT2 (HR 1.23; 95% CI 1.00, 1.55) (Figure 1). The estimates for 24-hour sleep duration suggested no causal effect on AMI per hour increase in sleep duration in UKBB (HR 0.97; 95% CI 0.75, 1.29) and HUNT2 (HR 0.76; 95% CI 0.31, 1.79). The sleep duration findings were further investigated using genetic variants specifically associated with short and long sleep duration. There was weak evidence for an adverse causal effect on AMI per doubling in odds of short sleep in UKBB (HR 1.14; 95% CI 0.97, 1.32) but not in HUNT2 (HR 0.87; 95% CI 0.15, 3.24). However, there was evidence for a protective causal effect on AMI per doubling in odds of long sleep in UKBB (HR 0.83; 95% CI 0.67, 0.99), which was underpowered in HUNT2 (HR 0.53; 95% CI 0.01, 8.28). Also, there was some evidence for an adverse causal effect on AMI per doubling in odds of morning chronotype in UKBB (HR 1.06; 95% CI 0.99, 1.11) (Figure 1).

**Figure 1:**
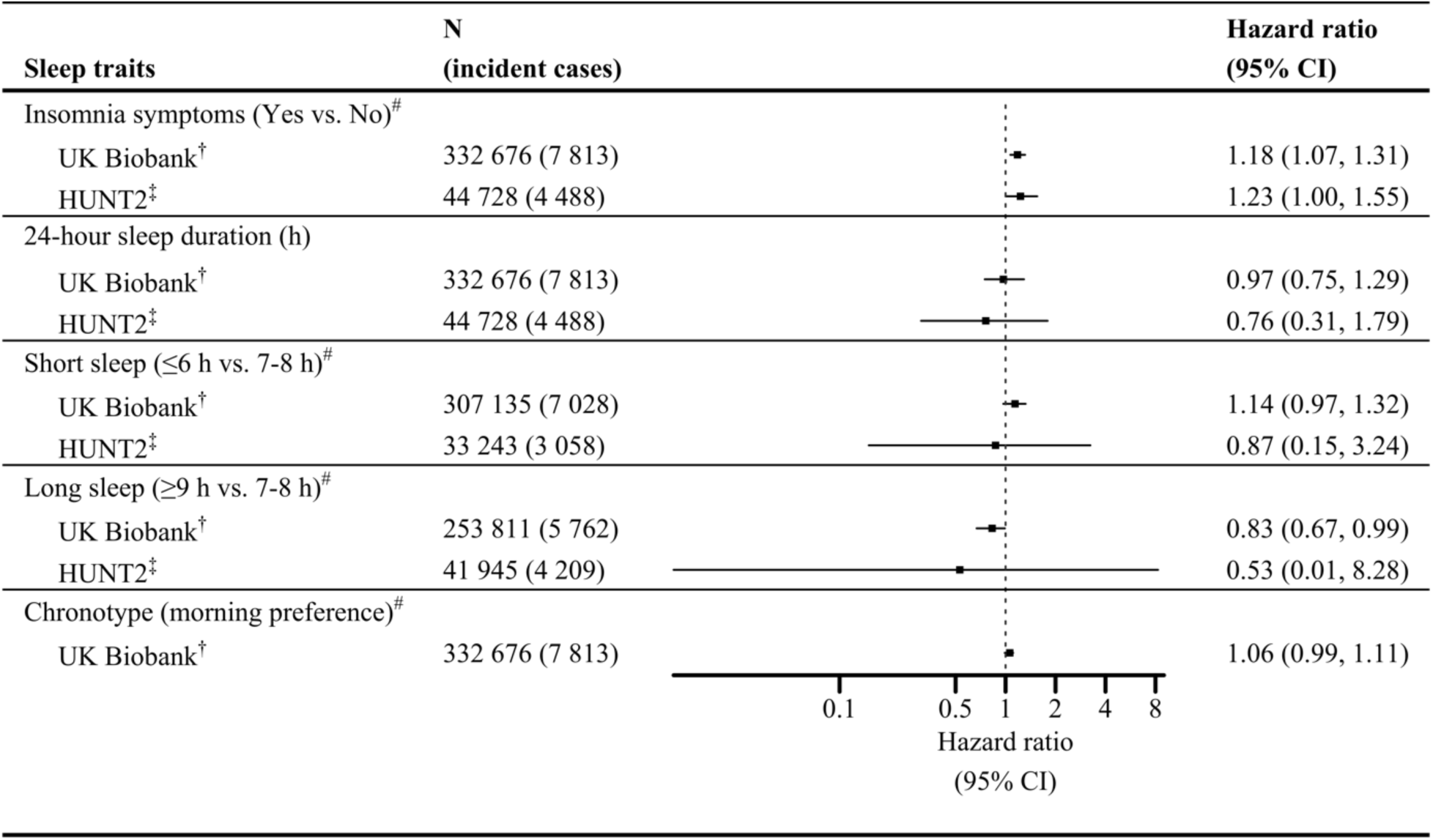
One-sample Mendelian randomization Cox regression analysis for risk of incident acute myocardial infarction associated with sleep traits in UK Biobank and HUNT2. CI indicates confidence interval. ^†^ Derived using unweighted genetic risk score for each sleep trait, with adjustment for age, gender, assessment centre, 40 genetic principal components, and genotyping chip. ^‡^ Derived using weighted genetic risk score for each sleep trait, with adjustment for age, gender, 20 genetic principal components, and genotyping chip. ^#^ Hazard ratio (95% CI) scaled to per doubling in odds of the sleep trait. Chronotype was missing in HUNT2.

### Joint causal effects of sleep traits on the risk of AMI

The distribution of the baseline characteristics across the factorial groups for any combinations of two sleep traits were equal (see Additional file 1: Tables S3 – S9), which indicates random allocation of the study participants into approximately equal-sized groups based on their genetic risk for any combinations of two sleep traits.

In UKBB, participants with high genetic risk for insomnia symptoms and high genetic risk for short sleep had slightly higher risk of AMI (HR 1.03; 95% CI 0.96, 1.10 and HR 1.05; 95% CI 0.98, 1.12, respectively), whereas participants with high genetic risk for both traits had the highest risk (HR 1.10; 95% CI 1.03, 1.12) (Figure 2), but there was no evidence of interaction (RERI 0.03; 95% CI −0.07, 0.12). This pattern was however not consistent in HUNT2, with imprecise estimates and a lack of evidence of interaction (RERI - 0.05; 95% CI −0.20, 0.09) (Figure 2). The joint effects of insomnia symptoms and long sleep on risk of AMI, were inconclusive in both UKBB and HUNT2 (Figure 2).

**Figure 2:**
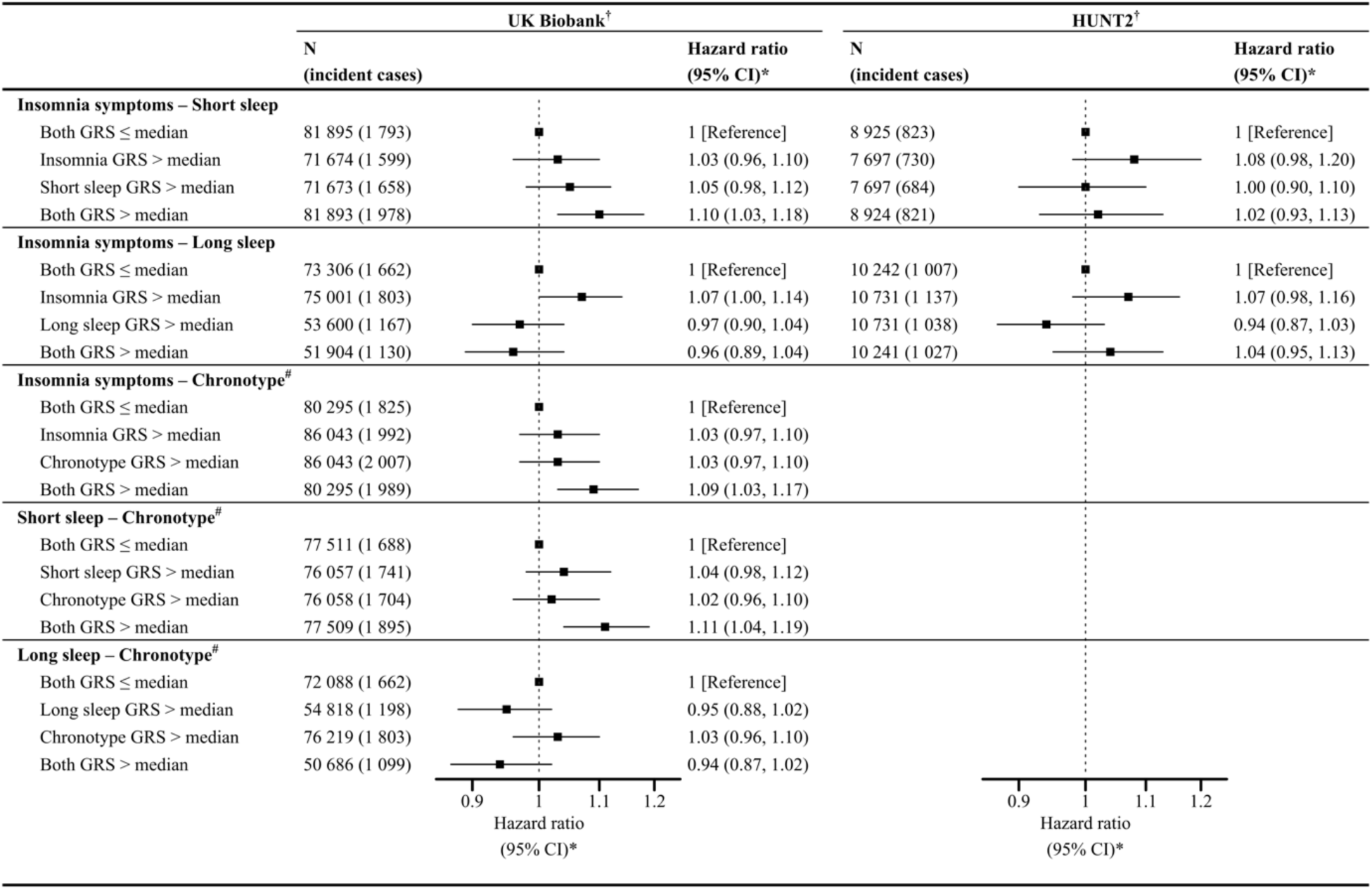
2×2 factorial Mendelian randomization Cox regression analysis assessing the joint effects of two sleep traits with risk of incident acute myocardial infarction in UK Biobank and HUNT2. CI indicates confidence interval; and GRS, genetic risk score. For each sleep trait combination, both GRS ≤ median represents low genetic risk for both sleep traits in combination, sleep trait 1 GRS > median represents high genetic risk for sleep trait 1 only, sleep trait 2 GRS > median represents high genetic risk for sleep trait 2 only and both GRS > median represents high genetic risk for both sleep traits. **^†^** Derived using unweighted genetic risk score for each sleep trait in UK Biobank, whereas using weighted genetic risk score for each sleep trait in HUNT2. **\*** Adjusted for age, gender, assessment centre (in UK Biobank), genetic principal components (40 in UK Biobank and 20 in HUNT2), and genotyping chip. **^#^** Chronotype genetic risk score calculated using alleles for morning preference.

In addition, UKBB participants with high genetic risk for insomnia symptoms and high genetic risk for a morning chronotype had slightly higher risk of AMI (HR 1.03; 95% CI 0.97, 1.10 and HR 1.03; 95% CI 0.97, 1.10, respectively) whereas participants with high genetic risk for both sleep traits had the highest risk (HR 1.09; 95% CI 1.03, 1.17) (Figure 2). There was no evidence of interaction (RERI 0.03; 95% CI −0.06, 0.12). Similarly, the UKBB participants with high genetic risk for short sleep and high genetic risk for a morning chronotype had slightly higher risk of AMI (HR 1.04; 95% CI 0.98, 1.12 and HR 1.02; 95% CI 0.96, 1.10, respectively), whereas participants with high genetic risk for both had the highest risk (HR 1.11; 95% CI 1.04, 1.19) (Figure 2), with no strong statistical evidence of interaction (RERI 0.05; 95% CI −0.05, 0.14). The joint effects of long sleep and morning chronotype were imprecise and not conclusive (Figure 2).

### Sensitivity analysis

The proportionality of hazards assumption was met for the one-sample and the 2×2 factorial MR Cox regression analyses (see Additional file 1: Tables S10 & S11).

The one-sample MR and 2×2 factorial MR estimates in HUNT2 using the uwGRS for the sleep traits remained unchanged (see Additional file 1: Table S12 & Figure S2).

After adjusting for multiple testing, several confounding factors were associated with the sleep trait uwGRS in UKBB, whereas only a few were associated with the sleep trait wGRS in HUNT2 (see Additional file 1: Tables S13 & S14). When the one-sample MR analysis adjusting for these potential confounding factors was carried out, evidence of adverse causal effects of insomnia symptoms was slightly weaker and less precise in UKBB (HR 1.04; 95% CI 0.92, 1.17) and HUNT2 (HR 1.13; 95% CI 0.87, 1.47) (see Additional file 1: Table S15).

The causal estimates obtained using MR-Egger, weighted median- and weighted mode-based methods attenuated slightly and were less precise (see Additional file 1: Figures S3-S7, Tables S16 & S17). The MR-Egger regression for insomnia symptoms in UKBB showed evidence of directional pleiotropy (HR 0.77; 95% CI 0.62, 0.95; and intercept 0.007; 95% CI 0.003, 0.012). Further, the post hoc one-sample MR analysis using insomnia symptoms variants from Lane *et al.* [24] gave similar estimates (see Additional file 1: Figure S8 and Table S18), where the MR-Egger regression showed evidence of directional pleiotropy in UKBB (HR 0.69; 95% CI 0.50, 0.96; and intercept 0.013; 95% CI 0.005, 0.022) and in HUNT2 (HR 0.97; 95% CI 0.78, 1.19; and intercept 0.006; 95% CI 0.001, 0.012).

The causal estimates were consistent when using GRS comprising 116 insomnia SNPs (one missing in HUNT imputed dataset) and 72 chronotype SNPs which replicated at genome-wide significance level (P <5×10^-8^) in the independent 23andMe dataset (see Additional file 1: Tables S19 & S20).

The estimates from the continuous factorial MR analysis using sleep trait GRS as quantitative traits (per SD increase) and their product term inferred similar effects (see Additional file 1: Figure S9). In UKBB, the GRS for insomnia symptoms and short sleep were independently linked to an increased risk of AMI (HR 1.03; 95% CI 1.01, 1.06 and HR 1.02; 95% CI 0.99, 1.04, respectively), with no evidence of interaction (RERI 0.02; 95% CI −0.01, 0.04). Similarly, the GRS for insomnia symptoms and morning chronotype were independently associated with an increased risk of AMI (HR 1.04; 95% CI 1.02, 1.06 and HR 1.03; 95% CI 1.00, 1.05, respectively), though there was no evidence of interaction (RERI 0.02; 95% CI −0.01, 0.04). Also, the GRS for short sleep and morning chronotype were both independently linked to an increased risk of AMI (HR 1.02; 95% CI 1.00, 1.04 and HR 1.03; 95% CI 1.00, 1.05, respectively), however suggested no evidence of interaction (RERI 0.01; 95% CI −0.02, 0.03).

On excluding the participants who self-reported the use of sleep medication(s), our one-sample and 2×2 factorial MR estimates remain unchanged (see Additional file 1: Figures S10 & S11).

## Discussion

Using individual-level data from the UKBB and HUNT2 cohorts, we performed one-sample and factorial MR analyses to investigate the causal effects of individual sleep traits (insomnia symptoms, sleep duration and morning chronotype) and their joint effects on the risk of AMI. We found evidence of an adverse causal effect of insomnia symptoms and a weak causal effect of short sleep on the risk of incident AMI, while long sleep had a protective effect in UKBB. We found no statistical evidence of interaction effects between sleep traits on the risk of AMI, but those with a high genetic risk for two sleep traits in combination (including insomnia symptoms, short sleep, and a morning chronotype) had the highest risk of AMI in UKBB. Moreover, our results showed a protective effect of genetically predisposed long sleep that was not affected by additionally being genetically predisposed to insomnia symptoms or a morning chronotype on incident AMI in UKBB. However, these results were not replicated in HUNT2, where the estimates were imprecise. These findings indicate that the main effects of sleep traits on the risk of AMI are likely to be independent of each other.

### Comparison with other studies

Direct comparison of MR results with observational findings is limited given that inherited genetic variation influences sleep behaviours over the life course, whereas observational estimates represent sleep behaviours measured at one time-point. Additionally, caution should be made when comparing our findings with other studies, due to variation in the definitions used for sleep traits.

#### Causal effects of individual sleep traits on the risk of AMI

Nonetheless, our finding showing evidence of an adverse causal effect of insomnia symptoms and a weak adverse causal effect of short sleep on the risk of AMI is consistent with prior observational [9, 11, 14] and MR research [11, 28, 29]. Our causal estimate of short sleep on the risk of AMI in UKBB was weaker compared to Daghlas *et al.* [11] (odds ratio (OR) 1.21; 95% CI 1.08, 1.37) and Ai *et al.* [29] (OR 1.21; 95% CI 1.09, 1.34), which might be due to different methodological approaches. Our analyses relied on survival data and reported HR considering incident cases of AMI on follow-up after recruitment in the cohorts, rather than OR. Our finding suggests a protective causal effect of long sleep on the risk of AMI contradicts with prior observational studies [11, 14], but aligns with a weak concordant effect shown by another MR study [29]. Long sleep may be an indicator of poor health status, being closely associated with depression, poor sleep quality, sedentary lifestyles and underlying comorbid conditions [63, 64], and so residual confounding or reverse causation may have biased previous observational findings. Moreover, our finding suggesting a weak causal effect of morning chronotype on the risk of AMI is inconsistent with our prior study that identified evening chronotype as detrimental [14]. It is likely that the previously reported protective association of morning chronotype is confounded.

#### Joint causal effects of sleep traits on the risk of AMI

Our finding that UKBB participants with high genetic risk for both insomnia symptoms and short sleep had the highest risk of AMI is consistent with evidence from our previous observational study where we found that insomnia symptoms and short sleep together increased the risk of AMI in UKBB more than the risk attributed to either insomnia symptoms or short sleep alone [14], and is supported by finding from another prospective study [16]. Moreover, our finding suggesting no interaction between insomnia symptoms and short sleep on risk of AMI is also in line with prior research [14, 16]. However, our finding of no positive interaction between insomnia symptoms and long sleep on the risk of AMI in UKBB contrasts with our previous observational study [14], where insomnia symptoms and long sleep together were found to increase the risk of AMI beyond their mere additive effects. This observed interaction could be due to confounding apparent in conventional observational studies, where poor health could be a confounder that would lead to false indications of harmful consequences of prolonged sleep. As previously mentioned, our finding in UKBB suggests a protective effect of genetic predisposition to long sleep on incident AMI, which was not affected by additionally being genetically predisposed to insomnia symptoms.

Our findings that UKBB participants with high genetic risk for both insomnia symptoms and a morning chronotype; and those with high genetic risk for both short sleep and a morning chronotype had the highest risk of AMI are in contrast with our observational study where we found evening chronotype to be more deleterious than morning chronotype in combination with insomnia symptoms or short sleep [14]. Although there was no interaction, these findings may suggest that the weak adverse effect of morning chronotype on AMI might partly be explained by concomitant genetic predisposition to insomnia symptoms or short sleep. Our finding that UKBB participants with high genetic risk for both long sleep and a morning chronotype likely decreased the risk of AMI is incongruous to our previous observational study, where long sleep together with morning chronotype was associated with an increased risk [14]. Again, there was no interaction and — if anything — our finding suggests a protective effect of genetic predisposition to long sleep on incident AMI, which was not affected by additionally being genetically predisposed to morning chronotype.

### Potential mechanisms

The underlying mechanisms by which insomnia symptoms or short sleep increase in the risk of AMI are multifactorial [65]. Insomnia and short sleep independently increase the risk of autonomic dysfunction, by increasing sympathetic tone (stress response) consequently accompanied by increased metabolic rate, increased heart rate and decreased heart rate variability [66–69]. Further, experimentally induced sleep restriction has been shown to cause hormonal imbalance which stimulate proinflammatory pathways [70], increase appetite [71, 72] and increase insulin resistance [73]. These autonomic and hormonal disturbances lead to hypertension [74, 75], diabetes [73], dyslipidaemia and obesity [71, 72], thus constituting a set of interrelated metabolic disorders that are pathophysiological in the development of cardiac dysfunction by accelerating endothelial dysfunction and atherosclerosis [76].

Our findings and these potential mechanisms might raise a concern that insomnia symptoms and short sleep could be regarded as similar traits. However, insomnia symptoms and sleep duration were found only moderately phenotypically (r = −0.25; P <0.001) and genetically (r_g_ = −0.50; P <6×10^-17^) correlated to each other [77]. It is also important to highlight that our findings on the joint causal effects of insomnia symptoms and short sleep on the risk of AMI do not employ that concomitant presence of insomnia symptoms and short sleep causes higher increase in risk of AMI through overstimulation of the suggested underlying mechanisms, or involve any supplementary mechanisms yet to be determined.

The underlying mechanism by which chronotype may influence AMI is not yet established. Studies have found evening chronotypes have more susceptibility for cardiometabolic risk behaviours and risk factors [12, 78, 79]. On the contrary, our causal findings suggesting that having a morning chronotype may be detrimental for incident AMI compared to having an evening chronotype, might be explained by the concomitant genetic predisposition to insomnia symptoms or short sleep.

### Strengths and limitations

This MR study leverages genetic information to assess the causal relationships between sleep traits and AMI, reducing the potential bias due to residual confounding, reverse causation and measurement error in conventional observational studies [22]. The novelty of this study is our application of factorial MR to explore the causal interplay between sleep traits on the risk of AMI, where participants were grouped based on their genetic predisposition for multiple sleep traits [60]. We are not aware of another study that has investigated the joint effects of sleep traits in the MR context. Another major novelty is that the study benefitted from the use of results from three large GWASs for insomnia symptoms [30], sleep duration [31] and chronotype [32], and used two large cohorts (UKBB and HUNT2) to replicate the findings. Moreover, this study draws on the principle of triangulation [80], where findings were compared from different methodological approaches, which further strengthened evidence supporting causation.

Nonetheless, there are a number of limitations of this study. Factorial MR analysis is usually underpowered to detect interaction which may raise the concerns of false negative results [60]. However, this study included the UKBB cohort with 332 676 participants constituting the largest factorial MR study on sleep traits to date. The strong instrument strength observed in UKBB cohort partially overcomes concerns due to underpowered factorial MR findings [81]. Another limitation is that although factorial MR can identify whether two independent exposures interact and have a joint effect of public health importance [81], it assumes exposures remain stable throughout the life course. Thus, the magnitude of effects should be cautiously interpreted.

Also, the validity of MR findings can be weakened by pleiotropy [82]. We used several sensitivity analyses to investigate possible sources of bias in MR. We found that the genetic risk for insomnia symptoms was strongly associated with BMI, smoking status, depression, and education among other covariates [30], which may be indicative of confounding, mediation or horizontal pleiotropy. Further to this, our results remained consistent across various MR methods, except for insomnia symptoms which showed evidence of an unbalanced pleiotropy in MR-Egger analysis. Additionally, previous studies have shown only mild attenuation of causal effects of insomnia symptoms on CAD risk when adjusted for BMI, smoking, depression and education using multivariable Mendelian randomization (MVMR) [25, 26]. Moreover, simulations have shown that MR-Egger may be unreliable when applied to a single dataset [59], and this is a limitation of our study.

The sleep traits were based on self-report. It remains unclear if self-reported sleep duration represents time in bed or actual sleep time. Also, the insomnia questions in UKBB or HUNT2 did not cover all aspects of insomnia (difficulty falling asleep, night awakenings, waking up early and daytime impairments) [83]. Chronotype in this study was assessed from a single question in UKBB, whereas validated instruments such as the Morningness-Eveningness Questionnaire and the Munich Chronotype Questionnaire use diverse questions to better estimate chronotype [84, 85]. Other sleep traits (e.g., sleep apnea, snoring, daytime napping) were not included, and we do not know whether these interact with insomnia symptoms or sleep duration. Moreover, the sleep traits we used are binary exposures (except for 24-hour sleep duration), which are likely coarsened approximations of the true latent exposure [86]. This opens up alternate pathways from the genetic instruments to the outcome, which may violate the exclusion restriction assumption, resulting in biased effect estimates [86]. In addition, causal estimates from MR of binary exposures on a binary outcome are difficult to interpret [87].

Due to the small sample size in HUNT2, we might have missed weak causal effects due to insufficient power. In addition, the genetic instrument explained little variance in short sleep and long sleep within HUNT2, implying possible weak instrument bias [88] and leading to wide CIs as shown in the bootstrap simulations [89]. Furthermore, SNPs for short and long sleep were not replicated in other independent cohorts [31], meaning that the GRS used is not validated in any other population.

The inclusion of UKBB in all exposure GWASs could lead to winner’s curse, that might bias the causal estimates in UKBB [90]. We therefore used unweighted GRS for our exposures in UKBB as recommended [51]. Also, we derived GRS for insomnia symptoms and chronotype composed of SNPs that replicated in an independent study (23andMe) [30, 32], which showed similar estimates, indicating winner’s curse is unlikely to have substantially biased effect estimates. However, we could not apply the same approach to explore the impact of winner’s curse on the sleep duration due to the limited sample size of the replication datasets in those studies [31], meaning that genetic associations might be imprecise.

The variation in the occurrence of AMI between UKBB (2.35%) and HUNT2 (10.03%) may be attributed to several factors related to the composition of the cohorts: a) the HUNT2 cohort followed up relative older participants, aged 20 years or above, with a mean baseline age of 48 years, while UKBB consisted of participants aged 40 to 69 years, with a mean baseline age of 56 years; b) the duration of follow-up was longer in HUNT2, spanning 20.4 years, compared to UKBB’s follow-up period of 11.7 years; c) UKBB (5.5% response rate) may represent a healthier sample [91], whereas HUNT2 (69.5% response rate) may be a more representative sample [36]; and d) baseline differences in the two underlying populations or differences due to time trend (for example, more current smokers in HUNT2 which was conducted about a decade earlier than UKBB). Moreover, competing risk from death among participants would potentially hinder the occurrence of AMI, that might overestimate the risks [92]. This is another limitation of our study.

Finally, our findings rely on analyses in UKBB due to its large sample. However, the generalizability of these findings may be limited due to a selected sample (5.5% response rate) in the UKBB cohort, which can bias both observational and MR estimates [93, 94]. Selection bias may artificially induce associations between genetic variants and confounders leading to the instrumental variable becoming invalid [95]. This might partly explain differences in UKBB and HUNT2 estimates observed in this study, where HUNT2 sample (69.5% response rate) more closely represents target population. The difference in demographics of the two cohorts might also cause inconsistent estimates. Moreover, the inclusion of cohorts from the European ancestry may further restrict generalizability of our findings.

## Conclusions

This study reveals no interaction effects between sleep traits on the risk of AMI, but found that two sleep traits in combination (including insomnia symptoms, short sleep, and a morning chronotype) had the highest risk of AMI. The role of chronotype in AMI risk remains uncertain, as the adverse causal effect of morning chronotype could partly be explained by genetic predisposition to insomnia symptoms or short sleep. This indicates that the main effects of insomnia symptoms and short sleep are likely to be independent of each other, i.e., the magnitude of the effect of insomnia symptoms on AMI does not depend on whether there is accompanying genetic predisposition to short sleep, and vice-versa. Thus, interventions targeting both insomnia symptoms and short sleep could be relevant for preventive initiatives to reduce the risk of AMI. Moreover, this study also suggests a potential protective effect of genetically predisposed long sleep that was not affected by additionally being genetically predisposed to insomnia symptoms and a morning chronotype.

## Supplementary material

Supplementary material is available online.

## Description of additional files

**Additional file 1:** *Information on covariates*; *Supplementary figures* **Figure S1.** Flow chart of the participant selection process. **Figure S2.** 2×2 factorial Mendelian randomization Cox regression analysis assessing the joint effects of two sleep traits with risk of incident acute myocardial infarction in HUNT2 using weighted and unweighted genetic risk scores for sleep traits. **Figure S3.** Association of insomnia SNPs from Jansen *et al*., 2019 and acute myocardial infarction (AMI) within a) UK Biobank b) HUNT2. IVW, MR-Egger, simple median and weighted median estimates are indicated by the red, green, blue and purple lines respectively. **Figure S4.** Association of 24-hour sleep duration SNPs from Dashti *et al*., 2019 and acute myocardial infarction (AMI) within a) UK Biobank b) HUNT2. IVW, MR-Egger, simple median and weighted median estimates are indicated by the red, green, blue and purple lines respectively. **Figure S5.** Association of short sleep duration SNPs from Dashti *et al*., 2019 and acute myocardial infarction (AMI) within a) UK Biobank b) HUNT2. IVW, MR-Egger, simple median and weighted median estimates are indicated by the red, green, blue and purple lines respectively. **Figure S6.** Association of long sleep duration SNPs from Dashti *et al*., 2019 and acute myocardial infarction (AMI) within a) UK Biobank b) HUNT2. IVW, MR-Egger, simple median and weighted median estimates are indicated by the red, green, blue and purple lines respectively. **Figure S7.** Association of chronotype (morning preference) SNPs from Jones *et al*., 2019 and acute myocardial infarction (AMI) within UK Biobank. IVW, MR-Egger, simple median and weighted median estimates are indicated by the red, green, blue and purple lines respectively. **Figure S8.** Association of insomnia SNPs from Lane *et al*., 2019 and acute myocardial infarction (AMI) within a) UK Biobank b) HUNT2. IVW, MR-Egger, simple median and weighted median estimates are indicated by the red, green, blue and purple lines respectively. **Figure S9.** Continuous factorial Mendelian randomization analysis using genetic risk score as quantitative traits with their product term assessing the joint effects of two sleep traits with risk of incident acute myocardial infarction in UK Biobank and HUNT2. **Figure S10.** One-sample Mendelian randomization Cox regression analysis for risk of incident acute myocardial infarction associated with sleep traits in UK Biobank and HUNT2 after excluding participants who reported self-reported use of sleep medication. **Figure S11.** 2×2 factorial Mendelian randomization Cox regression analysis assessing the joint effects of two sleep traits with risk of incident acute myocardial infarction in UK Biobank and HUNT2 after excluding participants who reported self-reported use of sleep medication; and *Supplementary tables* **Table S1.** Detailed summary of Mendelian randomization (MR) studies previously conducted on sleep traits and risk of coronary artery disease (CAD) or acute myocardial infarction (AMI). **Table S2.** Summary of genetic instruments showing their strength applying to UK Biobank and HUNT2. **Table S3.** Baseline characteristics of participants across groups categorized by dichotomizing to the median genetic risk scores for insomnia symptoms and short sleep in UK Biobank. **Table S4.** Baseline characteristics of participants across groups categorized by dichotomizing to the median genetic risk scores for insomnia symptoms and short sleep in HUNT2. **Table S5.** Baseline characteristics of participants across groups categorized by dichotomizing to the median genetic risk scores for insomnia symptoms and long sleep in UK Biobank. **Table S6.** Baseline characteristics of participants across groups categorized by dichotomizing to the median genetic risk scores for insomnia symptoms and long sleep in HUNT2. **Table S7.** Baseline characteristics of participants across groups categorized by dichotomizing to the median genetic risk scores for insomnia symptoms and chronotype (morning preference) in UK Biobank. **Table S8.** Baseline characteristics of participants across groups categorized by dichotomizing to the median genetic risk scores for short sleep and chronotype (morning preference) in UK Biobank. **Table S9.** Baseline characteristics of participants across groups categorized by dichotomizing to the median genetic risk scores for long sleep and chronotype (morning preference) in UK Biobank. **Table S10.** Statistical test of the proportional hazard assumption for one-sample Mendelian randomization (MR) Cox regression models. **Table S11.** Statistical test of the proportional hazard assumption for 2×2 factorial Mendelian randomization (MR) Cox regression models. **Table S12.** One-sample Mendelian randomization Cox regression analysis for risk of incident acute myocardial infarction associated with sleep traits in HUNT2 using weighted and unweighted genetic risk scores for sleep traits. **Table S13.** Associations between genetic risk scores and potential confounders in UK Biobank. **Table S14.** Associations between genetic risk scores and potential confounders in HUNT2. **Table S15.** One-sample Mendelian randomization analysis for risk of incident acute myocardial infarction associated with sleep traits with and without adjustment for potential confounders in UK Biobank and HUNT2. **Table S16.** Sensitivity analysis for risk of incident acute myocardial infarction associated with sleep traits in UK Biobank. **Table S17.** Sensitivity analysis for risk of incident acute myocardial infarction associated with sleep traits in HUNT2. **Table S18.** One-sample Mendelian randomization Cox regression analysis for risk of incident acute myocardial infarction associated with insomnia symptoms using instruments from Lane *et al*., 2019 in UK Biobank and HUNT2. **Table S19.** Sensitivity analysis for risk of incident acute myocardial infarction associated with insomnia symptoms and chronotype in UK Biobank using genetic variants genome-wide significant in 23andMe. **Table S20.** Sensitivity analysis for risk of incident acute myocardial infarction associated with insomnia symptoms in HUNT2 using genetic variants genome-wide significant in 23andMe. **Table S21.** List of medications used to define the sleep medication covariate in UK Biobank.

**Additional file 2:** *Genetic variants* **Table G1.** Summary information of genetic variants identified for insomnia symptoms. **Table G2.** Summary information of genetic variants identified for sleep duration. **Table G3.** Summary information of genetic variants identified for short sleep. **Table G4.** Summary information of genetic variants identified for long sleep. **Table G5.** Summary information of genetic variants identified for chronotype.

## Declarations

## Ethical approval and consent

UK Biobank received ethical approval from the National Health Service (NHS) Research Ethics Service (reference number 11/NW/0382). The HUNT Study was approved by the Data Inspectorate of Norway and recommended by the Regional Committee for Ethics in Medical Research (REK; reference number 152/95/AH/JGE). The ethical approval for conducting this study was also obtained from the Regional Committee for Ethics in Medical Research (REK nord; reference number 2020/47206).

Informed consent was obtained from all individual participants of both the cohorts included in this study.

## Availability of data and materials

The data supporting the findings are available in the supplementary material and upon request. The UK Biobank data is available to researchers, subject to successful registration and application process via their website (https://www.ukbiobank.ac.uk/). The data from the HUNT Study are available from the HUNT Research Centre but restrictions apply to the availability of these data, which were used under license for the current study, and so are not publicly available. However, the data are available for export given approval of application to the HUNT Research Centre (http://www.ntnu.edu/hunt/data). The data on hospital records linkages to the HUNT Study participants are available from Nord-Trøndelag Hospital Trust and require permission. All other data used are publicly available and referenced according in the main text.

## Abbreviations

AMI: Acute myocardial infarction
BMI: Body mass index
CAD: Coronary artery disease
CI: Confidence interval
GRS: Genetic risk score
GWAS: Genome-wide association study
HADS: Hospital Anxiety and Depression Scale
HES: Hospital Episode Statistics
HR: Hazard ratio
HUNT: The Trøndelag Health Study
ICD: International Classification of Diseases
MR: Mendelian randomization
MVMR: Multivariable Mendelian randomization
NHS: National Health Service
OR: Odds ratio
PEDW: Patient Episode Database for Wales
RERI: Relative excess risk due to interaction
SBP: Systolic blood pressure
SD: Standard deviation
SMR: Scottish Morbidity Record
SNP: Single nucleotide polymorphism
TDI: Townsend deprivation index
TSPS: Two-stage predictor substitution
UKBB: UK Biobank

## Acknowledgements

This research has been conducted using data from UK Biobank, a major biomedical database (www.ukbiobank.ac.uk), under application number 40135.

The Trøndelag Health Study (HUNT) is a collaboration between HUNT Research Centre (Faculty of Medicine and Health Sciences, Norwegian University of Science and Technology (NTNU)), Trøndelag County Council, Central Norway Regional Health Authority, and the Norwegian Institute of Public Health. We want to thank clinicians and other employees at Nord-Trøndelag Hospital Trust for their support and for contributing to data collection in this research project.

## Authors’ contributions

N.A. interpreted and analysed the data, interpreted the findings, and wrote the paper; L.B.S., and R.C.R. had the original idea for this study, interpreted the data, and critically revised the paper; E.S.S., B.M.B., and B.O.Å. had the original idea for this study, and critically revised the paper; L.B. assisted with analysis, and critically revised the paper; and H.D. assisted with interpreting data on acute myocardial infarction from medical records assessed through hospitals in the Trøndelag County, and critically revised the paper. All authors read and approved the final manuscript.

## Authors’ Twitter handles

Nikhil Arora: @dr_nikhil_arora

Laxmi Bhatta: @laxmibhatta001

Eivind Schjelderup Skarpsno: @E_Skarpsno Bjørn Olav Åsvold: @BAsvold

Ben Michael Brumpton: @bmbrumpton

Rebecca Claire Richmond: @BeckyRichmond90 Linn Beate Strand: @strandlb

