## Additional file 1 for "Investigating the causal interplay between sleep traits and risk of acute myocardial infarction: a Mendelian randomization study"

### Supplementary material

#### Information on covariates

A self-administered questionnaire was used to gather information on various covariates including gender, age at recruitment, marital status, alcohol intake, smoking status, physical activity, Townsend Deprivation Index (in UK Biobank only), education, shift work, and use of sleep medication(s). Additionally, participants attended examination stations where clinical examination was performed, and blood samples were drawn by trained staff.

In UKBB, participants were categorized as "Married" if they live with their husband/wife/partner, and as "Unmarried" if they don't. Also, the information about the number of people living in a household was used to categorize individuals living alone as "Unmarried" in cases where the marital status information was missing. In HUNT2, marital status was categorized into "Unmarried", "Married", and "Separated/Divorced/Widowed".

In UKBB and HUNT2, participants were asked about their alcohol intake frequency and were categorized as follows: "Never/rarely" for non-drinkers or those who only drink on special occasions, "Monthly" for those who drink 1 - 3 times a month, "Weekly" for those who drink 1- 4 times a week or "Daily/almost daily" for those who drink more frequently. Additionally, in HUNT2, the information about participants who had never consumed alcohol was used to categorize them as "Never/rarely" for observations having missing information on alcohol intake frequency. Thus, this information will be categorized as "Never/rarely", "Monthly", "Weekly" or "Daily/almost daily" alcohol intake.

The information on smoking status was categorized as "Never", "Previous" or "Current" smoker for UKBB and HUNT2.

In UKBB, physical activity (PA) was evaluated using adapted questions from the validated short International Physical Activity Questionnaire (IPAQ) [1], following guidelines published for data processing by IPAQ [2]. IPAQ assessed total physical activity, including walking, moderate, and vigorous PA performed over the last 7 days. Participants were categorized into three mutually exclusive PA categories: "High" (≥1 h of moderate PA or ≥½ h of vigorous PA above basal level of activity on most days), "Moderate" (≥½ h of moderate PA above basal level of activity on most days) or "Low/inactive" (anything else) based on a standard scoring criteria [3], where approximately 5000 steps per day was considered as basal activity. In HUNT2, PA was classified based on self-reported leisure time light and hard PA during the past year. Light PA was defined as activity that did not cause sweating or shortness of breath, while hard PA was defined as activity that resulted in sweating or shortness of breath. Participants were instructed to include the commute to work as leisure time. The study participants were grouped into three mutually exclusive categories: "High" (defined by ≥1 h of hard PA regardless of light PA or ≥3 h of light PA with <1 h of hard PA), "Moderate" (defined by ≥3 h of light PA with no hard PA or <3 h of light PA with <1 h of hard PA), or "Low/inactive" (for anything else). This categorization strategy for PA was previously used by Brumpton *et al.* [4]. The reliability and validity of the questions on PA from HUNT2 have been reported to be acceptable for hard PA and poor for light PA [5].

In UKBB and HUNT2, education level was classified into three categories: "10 years or less" (for primary and lower secondary school education), "11-13 years" (for upper secondary school education), or "14 years or more" (for university/college education).

In UKBB, the Townsend Deprivation Index (TDI) was used as a measure for socioeconomic status to account for socioeconomic disparities and urban-rural mix within the UK. The index was created from census data on housing, employment, car availability and social class based on postal codes of participants, with higher values indicating a higher level of deprivation. The TDI has been validated for use in a UK-based population [6]. The HUNT2 sample was based on the population of the northern region of Trøndelag County in Norway, which is fairly representative of Norway regarding socioeconomic characteristics [7]. Thus, any potential socioeconomic differences would be largely captured by education attainment.

The UKBB participants were asked about working shift work or working night shifts separately. These responses were then combined to create a proxy variable, with the highest response category being used as the final value. This proxy variable was then dichotomized, with “Usually” or “Always” being classified as “Yes”, and all other responses as “No”. In HUNT2, working shifts/at night/on call was also dichotomized as “Yes” or “No”. Additionally, the information on current employment/work status from both UKBB and HUNT2 was used to categorize those without paid employment or who were self-employed as “No” for observations with missing information on working shifts/at night/on call.

In UKBB, the use of sleep medication(s) was ascertained by the self-reported use of medications from the list of sleep medications as used by Daghlas *et al*. [8], along with five other commonly used anxiolytics or sleep medications (list included in

Table S21). These responses were then dichotomized as “Yes” or “No” for the use of sleep medication(s). In the HUNT2 study, participants were asked about their use of anxiolytics or sleep medications in the last month and categorized as “Yes” if they reported daily or weekly intake, and “No” otherwise.

*Clinical examination*

Within UKBB, weight was measured using the Tanita BC-418MA body composition analyser to the nearest 0.1kg and height was measured using a Seca 202 height measure. Within HUNT2, weight was measured to the nearest 0.5kg and height was measured to the nearest 1cm. Participants in both UKBB and HUNT2 wore light clothes and no shoes during these measurements. The body mass index (BMI) was then calculated by dividing weight (in kg) by the square of height (in metre).

In UKBB, systolic and diastolic measurements of blood pressure were recorded automatedly (using Omron HEM-705 IT electronic blood pressure monitor) and/or manually (using manual sphygmomanometer). Two sets of measurements were taken with a one-minute interval and the average of these two were used in our analyses. In cases where automated readings were not available, the manual readings were used. In HUNT2, systolic and diastolic measurements of blood pressure were recorded automatedly (using a Dinamap 845XT (Critikon) sphygmomanometer based on oscillometry). Three sets of measurements were taken with a one-minute interval, and the average of second and third measurements were used in the analysis.

*Laboratory measurements*

For UKBB, a random (non-fasting) blood sample was collected from each participant in accordance with standard operating procedures for the UKBB. The samples were stored in refrigerators between 2 to 8 °C. The fasting time was recorded as the interval between last consumption of food or drink and the blood sample being taken. The samples were transferred to a central laboratory for storage and analyses on a daily basis. The serum samples were centrifuged for 10 minutes at 2000 RCF and the serum concentrations of glucose, total cholesterol, HDL-cholesterol, and triglycerides were analysed using a Beckman Coulter AU5800 automated analyser. Glucose was measured using hexokinase analysis, while total cholesterol, HDL-cholesterol and triglycerides were measured by CHO-POD analysis, enzyme immunoinhibition analysis and GPO-POD analysis, respectively [9].

For HUNT2, a random (non-fasting) blood sample was collected from each participant. The samples were then analysed at the Central Laboratory, Levanger Hospital, using a Hitachi 911 Autoanalyzer (Hitachi, Mito, Japan). The serum was separated from the blood by centrifugation within 2 hours of collection and stored in a refrigerator (4 °C). Time between the last meal and venepuncture was recorded. The samples were sent to the laboratory on the same day or within two to three days (for example on weekends). The serum concentrations of glucose, total cholesterol, HDL-cholesterol, and triglycerides were analysed applying reagents from Boehringer Mannheim (Mannheim, Germany). The day-to-day coefficients of variation were 1.3-2.0%, 1.3-1.9%, 2.4%, and 0.7-1.3%, respectively. The glucose was measured using an enzymatic hexokinase method, total cholesterol and HDL-cholesterol were measured by an enzymatic colorimetric cholesterol esterase method, and triglycerides were measured with an enzymatic colorimetric method [7].

*Depression and anxiety*

For UKBB, hospitals recorded ICD-10 codes - F40 and F41 for anxiety; and F32, F33, F34, F38 and F39 for depression were used to identify participants with anxiety or depression episodes. From this information, two binary proxy variables were created for anxiety and depression, each categorized as “Yes” or “No”.

For HUNT2, the Hospital Anxiety and Depression Scale (HADS) was used to assess the symptoms of anxiety and depression. The questionnaire consisted of 14 Likert-scaled items (7 each for anxiety and depression) having a four-point scale ranging from 0 (not at all) to 3 (very often). Responses were summed to provide scores each for anxiety and depression ranging from 0 to 21. A higher score indicates an increased likelihood of anxiety and depression [10]. The HADS does not include somatic items or items about sleep difficulties. This tool is useful for assessing the symptom severity due to anxiety and depression in both primary care and hospital settings [11], and its psychometric properties have been validated as part of the HUNT study [12].

#### Supplementary figures

*Figure S1: Flow chart of the participant selection process.*

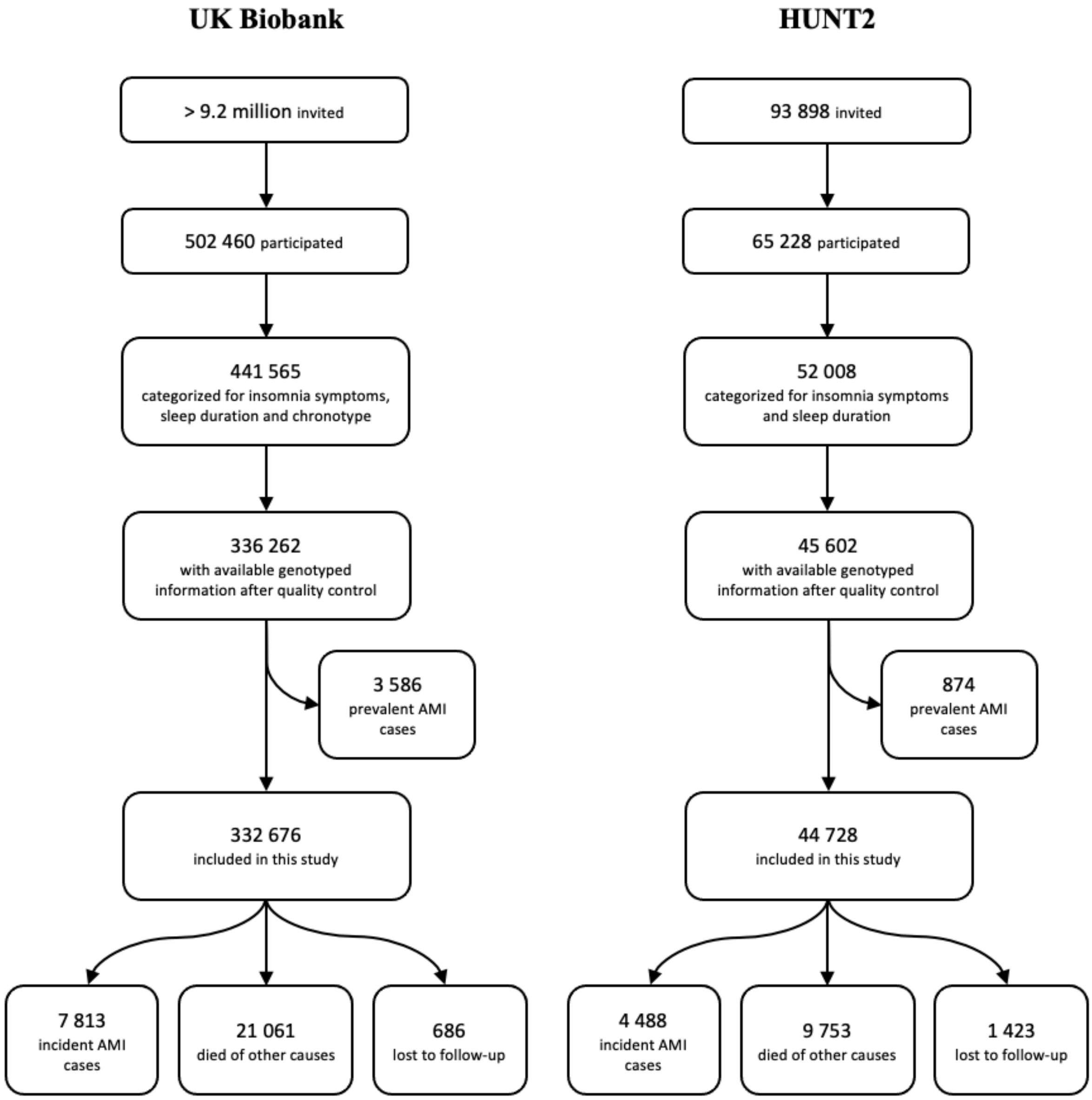

*Figure S2: 2x2 factorial Mendelian randomization Cox regression analysis assessing the joint effects of two sleep traits with risk of incident acute myocardial infarction in HUNT2 using weighted and unweighted genetic risk scores for sleep traits.*

| | Weighted genetic risk score | | Unweighted genetic risk score | |
|---|---|---|---|---|
| | **N (incident cases)** | **Hazard ratio (95% CI)*** | **N (incident cases)** | **Hazard ratio (95% CI)*** |
| **Insomnia symptoms – Short sleep** | | | | |
| Both GRS ≤ median | 8 925 (823) | 1 [Reference] | 8 797 (803) | 1 [Reference] |
| Insomnia GRS > median | 7 697 (730) | 1.08 (0.98, 1.20) | 7 830 (750) | 1.10 (1.00, 1.22) |
| Short sleep GRS > median | 7 697 (684) | 1.00 (0.90, 1.10) | 7 825 (708) | 1.04 (0.94, 1.14) |
| Both GRS > median | 8 924 (821) | 1.02 (0.93, 1.13) | 8 791 (797) | 1.03 (0.93, 1.13) |
| **Insomnia symptoms – Long sleep** | | | | |
| Both GRS ≤ median | 10 242 (1 007) | 1 [Reference] | 10 594 (1 029) | 1 [Reference] |
| Insomnia GRS > median | 10 731 (1 137) | 1.07 (0.98, 1.16) | 10 810 (1 137) | 1.10 (1.01, 1.20) |
| Long sleep GRS > median | 10 731 (1 038) | 0.94 (0.87, 1.03) | 10 380 (1 012) | 1.01 (0.92, 1.10) |
| Both GRS > median | 10 241 (1 027) | 1.04 (0.95, 1.13) | 10 161 (1 031) | 1.07 (0.98, 1.16) |

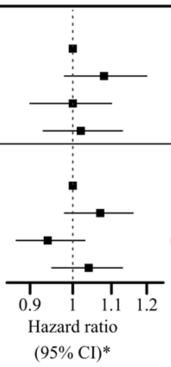


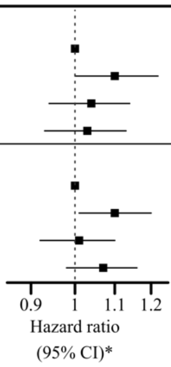


CI indicates confidence interval; and GRS, genetic risk score.
For each sleep trait combination, both GRS ≤ median represents low genetic risk for both sleep traits in combination, sleep trait 1 GRS > median represents high genetic risk for sleep trait 1 only, sleep trait 2 GRS > median represents high genetic risk for sleep trait 2 only and both GRS > median represents high genetic risk for both sleep traits.
* Adjusted for age, gender, 20 genetic principal components, and genotyping chip.

*Figure S3: Association of insomnia SNPs from Jansen et al., 2019 [13] and acute myocardial infarction (AMI) within a) UK Biobank b) HUNT2. IVW, MR-Egger, simple median and weighted median estimates are indicated by the red, green, blue and purple lines respectively.*

a)

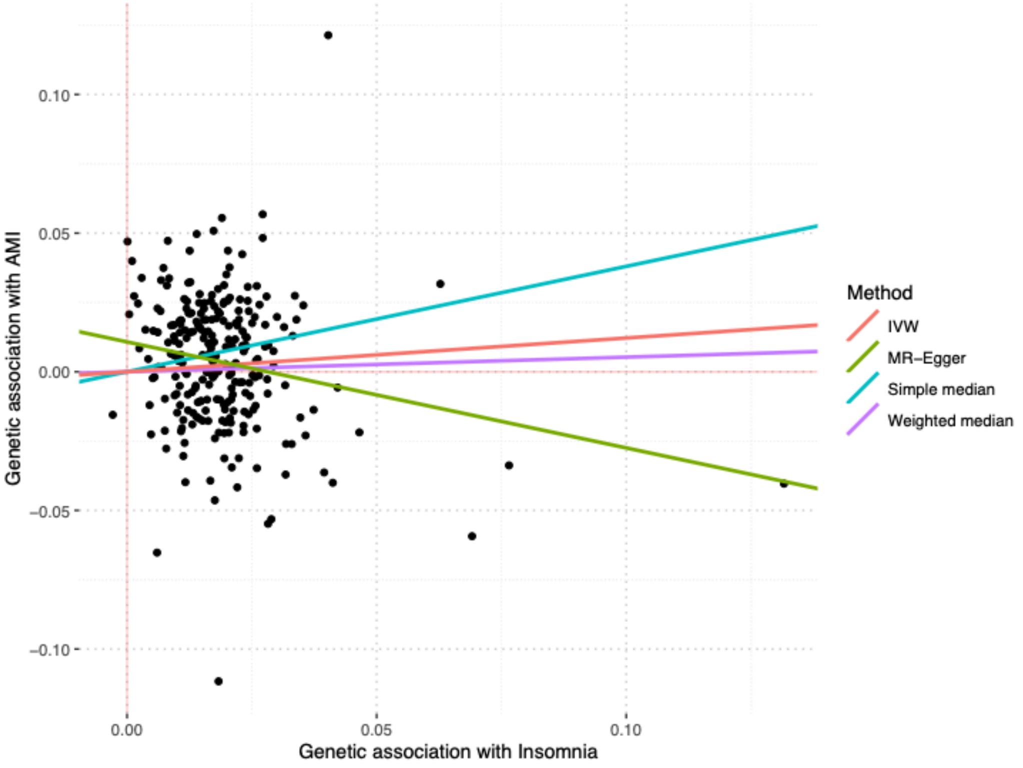


b)

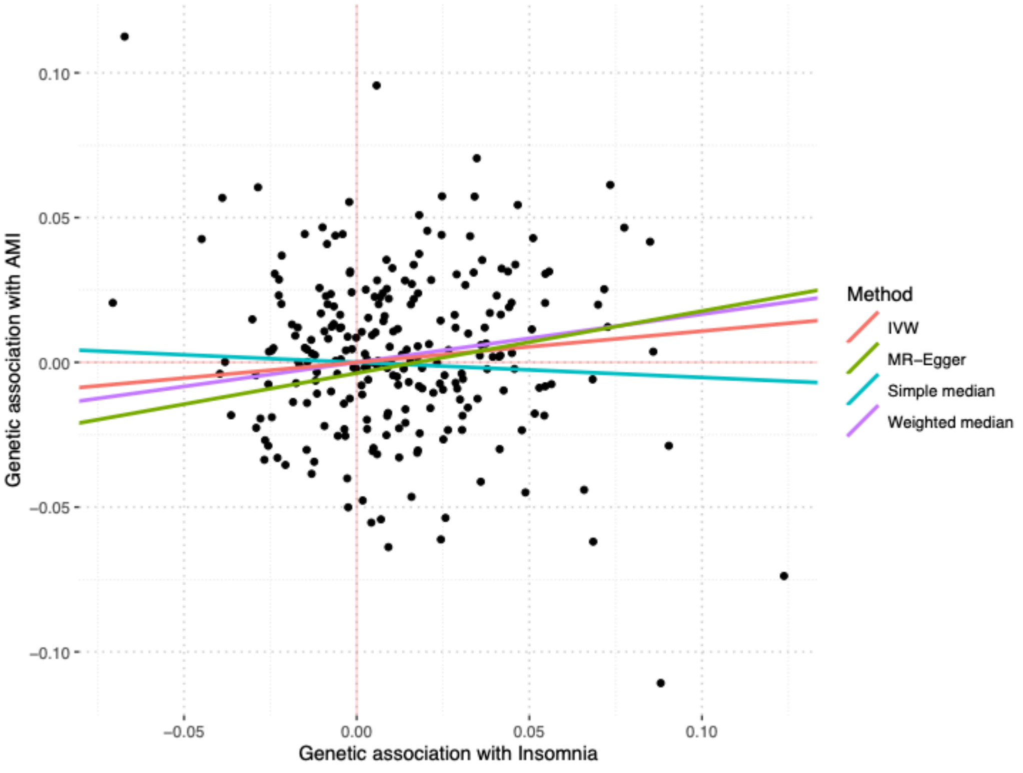

*Figure S4: Association of 24-hour sleep duration SNPs from Dashti et al., 2019 [14] and acute myocardial infarction (AMI) within a) UK Biobank b) HUNT2. IVW, MR-Egger, simple median and weighted median estimates are indicated by the red, green, blue and purple lines respectively.*

a)

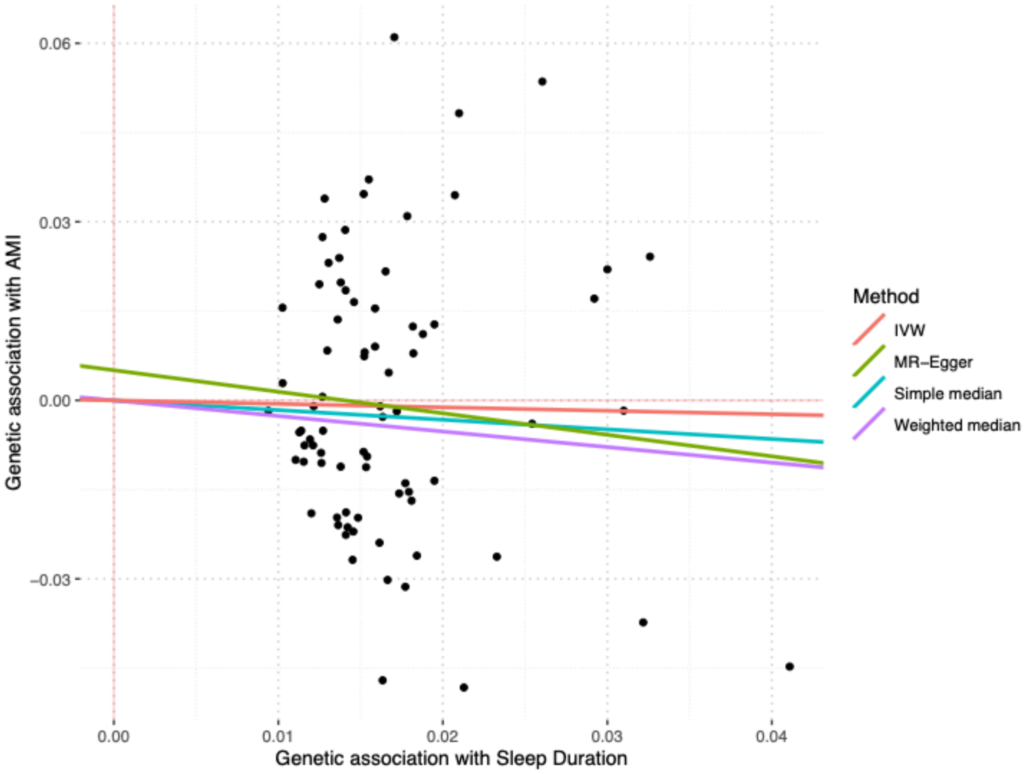


b)

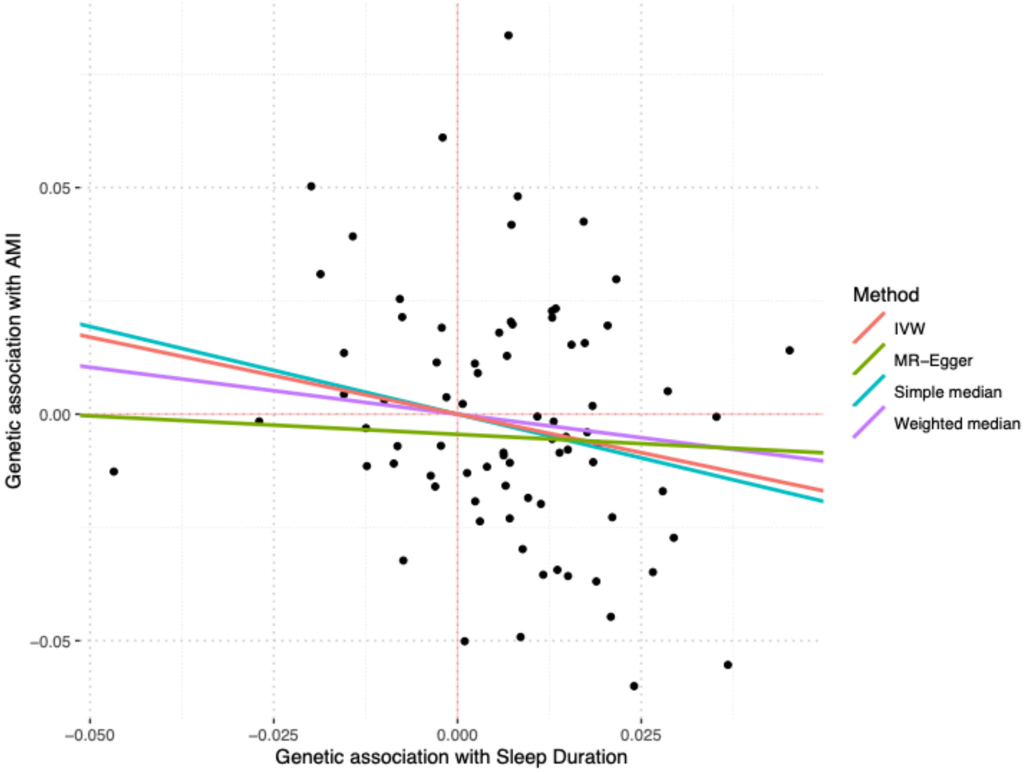

*Figure S5: Association of short sleep duration SNPs from Dashti et al., 2019 [14] and acute myocardial infarction (AMI) within a) UK Biobank b) HUNT2. IVW, MR-Egger, simple median and weighted median estimates are indicated by the red, green, blue and purple lines respectively.*

a)

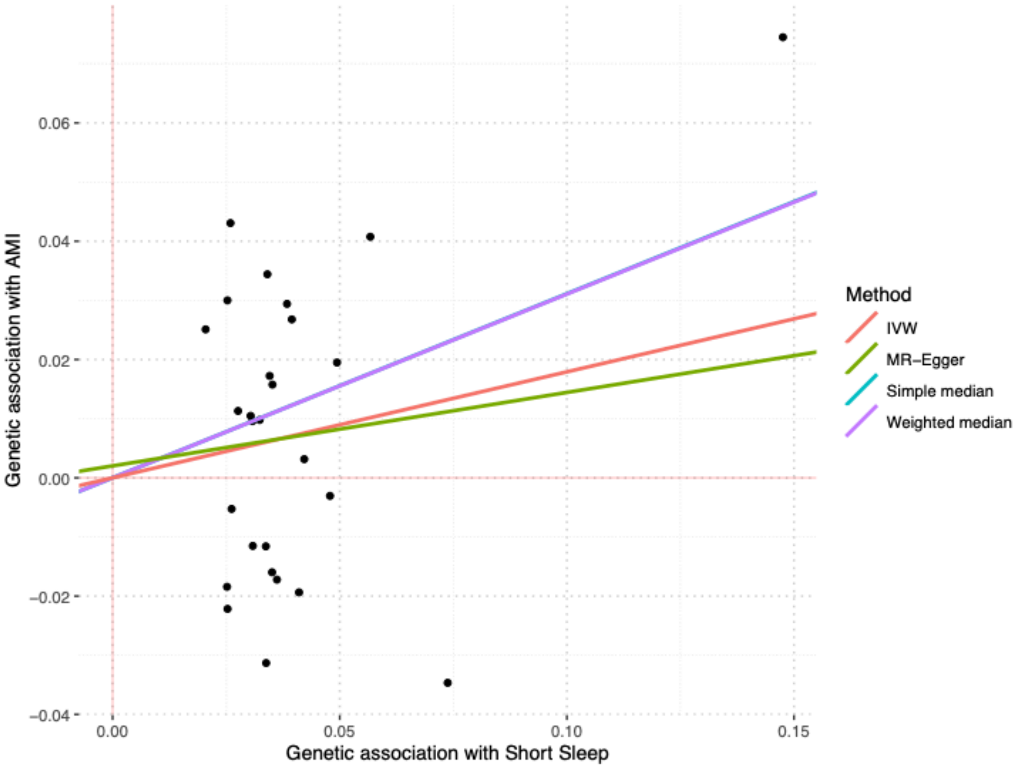


b)

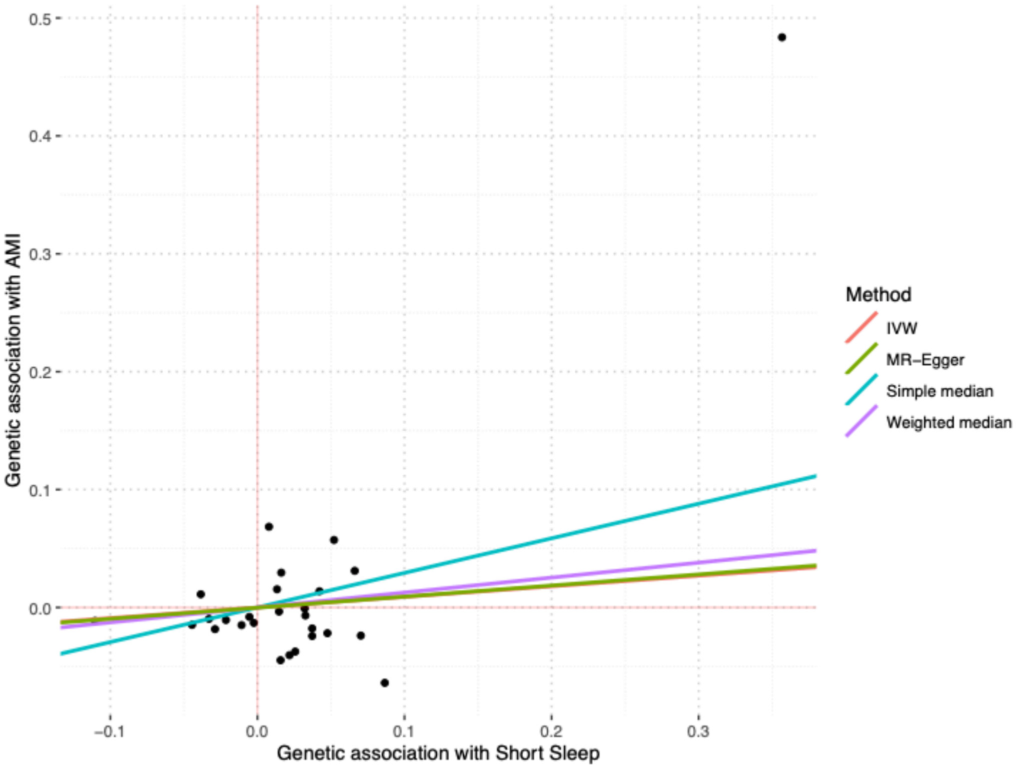

*Figure S6: Association of long sleep duration SNPs from Dashti et al., 2019 [14] and acute myocardial infarction (AMI) within a) UK Biobank b) HUNT2. IVW, MR-Egger, simple median and weighted median estimates are indicated by the red, green, blue and purple lines respectively.*

a)

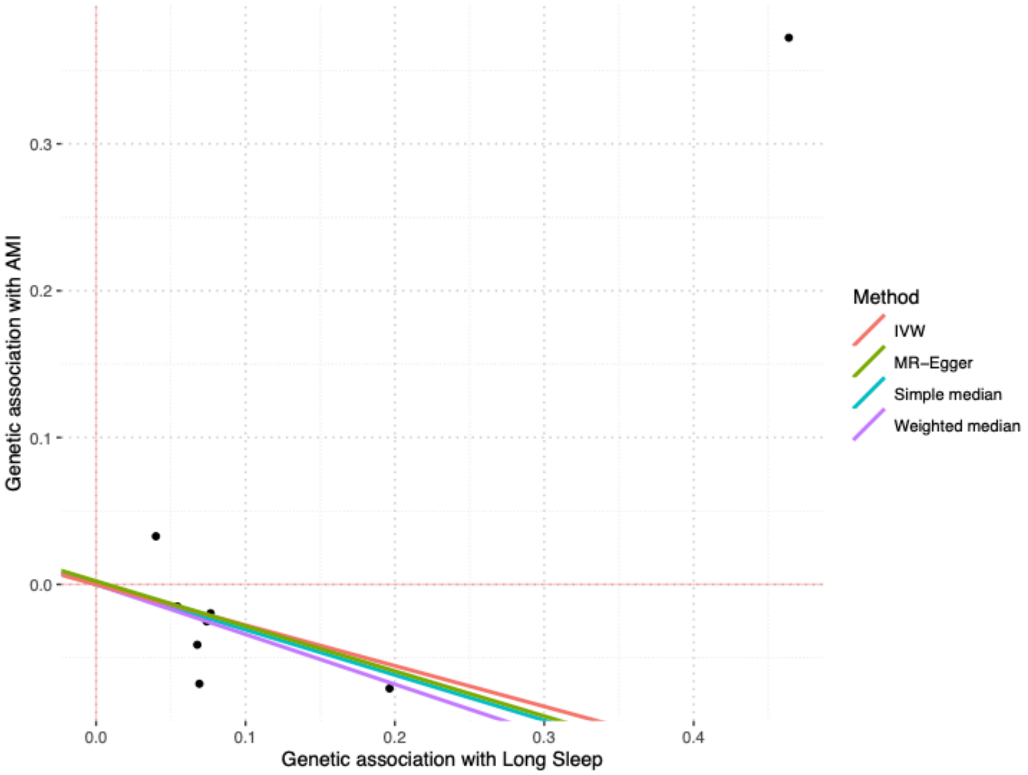


b)

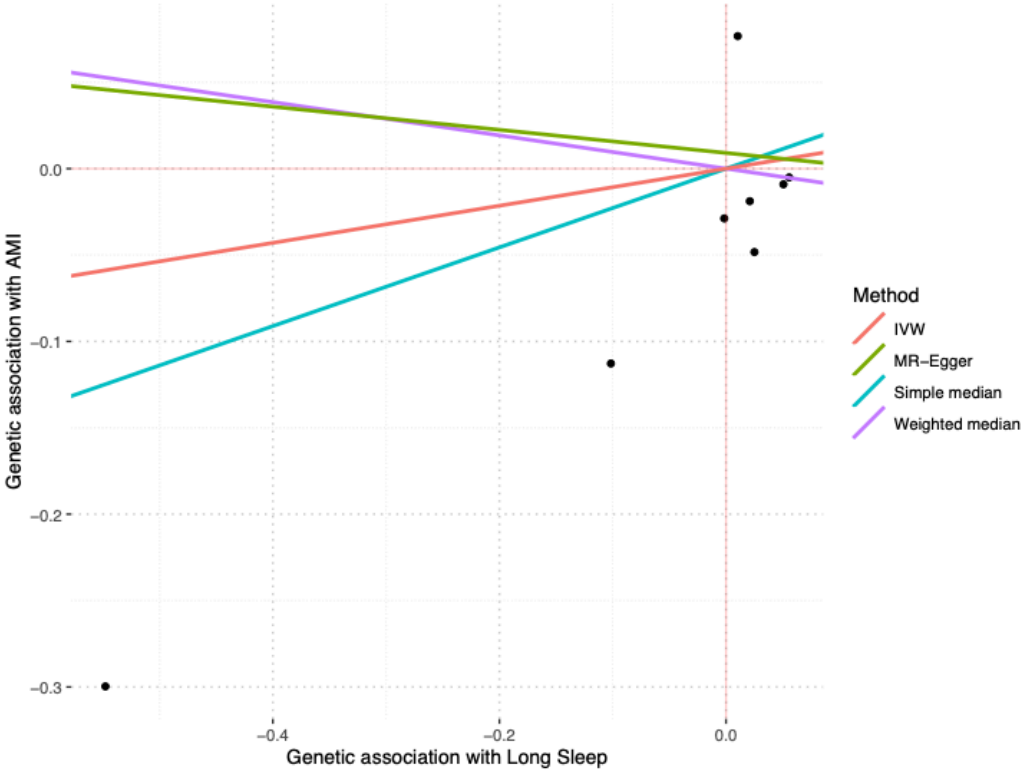

*Figure S7: Association of chronotype (morning preference) SNPs from Jones et al., 2019 [15] and acute myocardial infarction (AMI) within UK Biobank. IVW, MR-Egger, simple median and weighted median estimates are indicated by the red, green, blue and purple lines respectively.*

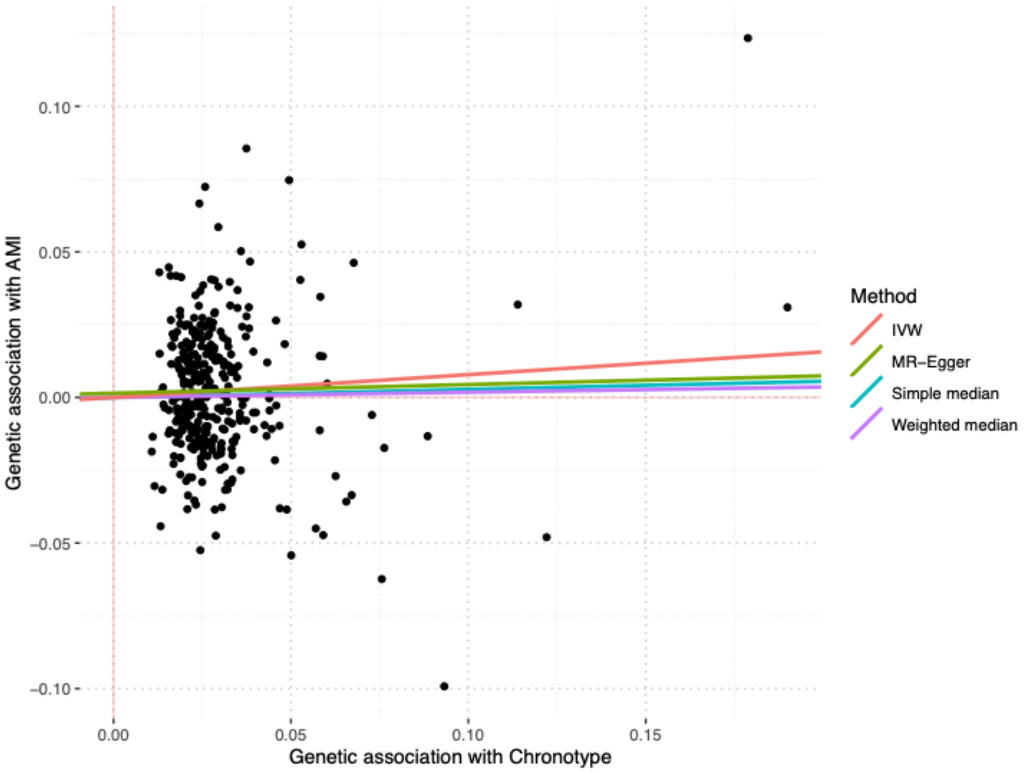

*Figure S8: Association of insomnia SNPs from Lane et al., 2019 [16] and acute myocardial infarction (AMI) within a) UK Biobank b) HUNT2. IVW, MR-Egger, simple median and weighted median estimates are indicated by the red, green, blue and purple lines respectively.*

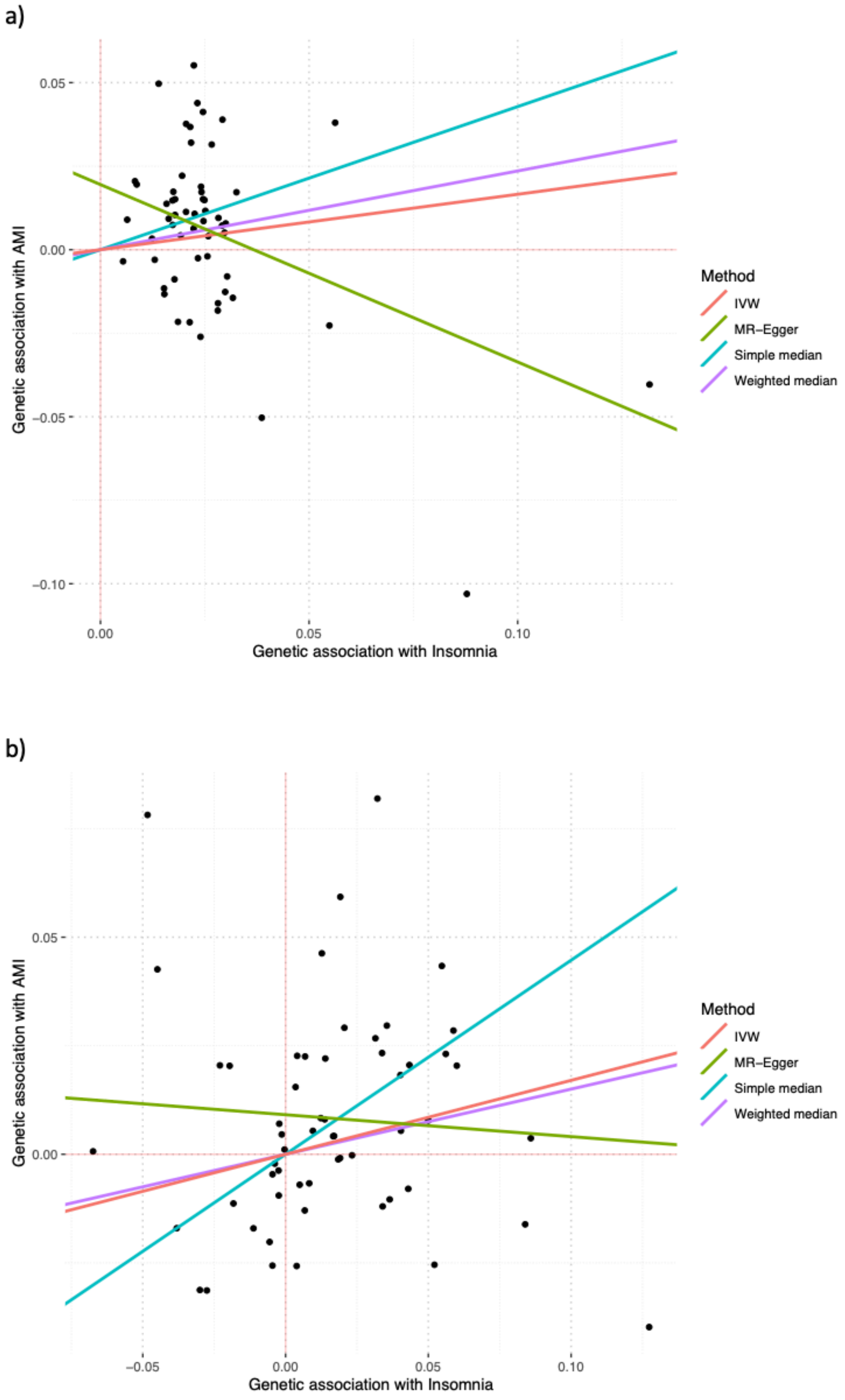

*Figure S9: Continuous factorial Mendelian randomization analysis using genetic risk score as quantitative traits with their product term assessing the joint effects of two sleep traits with risk of incident acute myocardial infarction in UK Biobank and HUNT2.*

| | UK Biobank[†] | | HUNT2[†] | |
|---|---|---|---|---|
| | N (incident cases) | Hazard ratio (95% CI)* | N (incident cases) | Hazard ratio (95% CI)* |
| **Insomnia symptoms – Short sleep** | | | | |
| Insomnia GRS (1SD increase) | 307 135 (7 028) | 1.03 (1.01, 1.06) | 33 243 (3 058) | 1.03 (0.99, 1.06) |
| Short sleep GRS (1SD increase) | 307 135 (7 028) | 1.02 (0.99, 1.04) | 33 243 (3 058) | 0.99 (0.95, 1.02) |
| Product term of two scores | | 1.02 (0.99, 1.04) | | 0.99 (0.95, 1.02) |
| **Insomnia symptoms – Long sleep** | | | | |
| Insomnia GRS (1SD increase) | 253 811 (5 762) | 1.03 (1.00, 1.06) | 41 945 (4 209) | 1.04 (1.01, 1.07) |
| Long sleep GRS (1SD increase) | 253 811 (5 762) | 0.98 (0.95, 1.00) | 41 945 (4 209) | 0.98 (0.95, 1.01) |
| Product term of two scores | | 0.99 (0.97, 1.02) | | 1.00 (0.97, 1.02) |
| **Insomnia symptoms – Chronotype[#]** | | | | |
| Insomnia GRS (1SD increase) | 332 676 (7 813) | 1.04 (1.02, 1.06) | | |
| Chronotype GRS (1SD increase) | 332 676 (7 813) | 1.03 (1.00, 1.05) | | |
| Product term of two scores | | 1.01 (0.99, 1.04) | | |
| **Short sleep – Chronotype[#]** | | | | |
| Short sleep GRS (1SD increase) | 307 135 (7 028) | 1.02 (1.00, 1.04) | | |
| Chronotype GRS (1SD increase) | 307 135 (7 028) | 1.03 (1.00, 1.05) | | |
| Product term of two scores | | 1.00 (0.98, 1.03) | | |
| **Long sleep – Chronotype[#]** | | | | |
| Long sleep GRS (1SD increase) | 253 811 (5 762) | 0.98 (0.95, 1.00) | | |
| Chronotype GRS (1SD increase) | 253 811 (5 762) | 1.01 (0.98, 1.04) | | |
| Product term of two scores | | 0.99 (0.97, 1.02) | | |

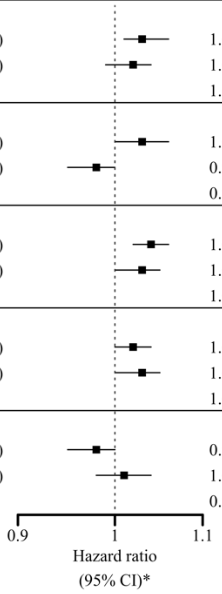


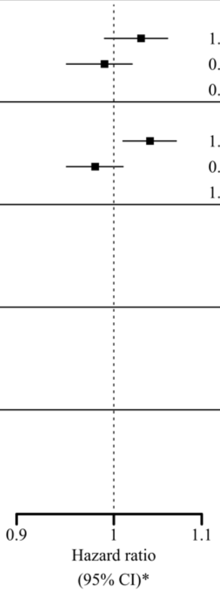


CI indicates confidence interval; GRS, genetic risk score; and SD, standard deviation
[†] Derived using unweighted genetic risk score for each sleep trait in UK Biobank, whereas using weighted genetic risk score for each sleep trait in HUNT2.
[*] Adjusted for age, gender, assessment centre (in UK Biobank), genetic principal components (40 in UK Biobank and 20 in HUNT2), and genotyping chip.
[#] Chronotype genetic risk score calculated using alleles for morning preference.

*Figure S10: One-sample Mendelian randomization Cox regression analysis for risk of incident acute myocardial infarction associated with sleep traits in UK Biobank and HUNT2 after excluding participants who reported self-reported use of sleep medication.*

| Sleep traits | N (incident cases) | Hazard ratio (95% CI) |
|---|---|---|
| Insomnia symptoms (Yes vs. No)[#] | | |
| UK Biobank[†] | 329 434 (7 706) | 1.17 (1.06, 1.29) |
| HUNT2[‡] | 41 785 (3 999) | 1.21 (0.97, 1.49) |
| 24-hour sleep duration (h) | | |
| UK Biobank[†] | 329 434 (7 706) | 0.97 (0.74, 1.28) |
| HUNT2[‡] | 41 785 (3 999) | 0.71 (0.30, 1.68) |
| Short sleep (≤6 h vs. 7-8 h)[#] | | |
| UK Biobank[†] | 304 345 (6 933) | 1.14 (0.98, 1.32) |
| HUNT2[‡] | 31 609 (2 797) | 0.98 (0.42, 2.30) |
| Long sleep (≥9 h vs. 7-8 h)[#] | | |
| UK Biobank[†] | 251 860 (5 707) | 0.83 (0.68, 1.00) |
| HUNT2[‡] | 39 167 (3 748) | 0.51 (0.18, 1.45) |
| Chronotype (morning preference)[#] | | |
| UK Biobank[†] | 329 434 (7 706) | 1.06 (0.99, 1.11) |

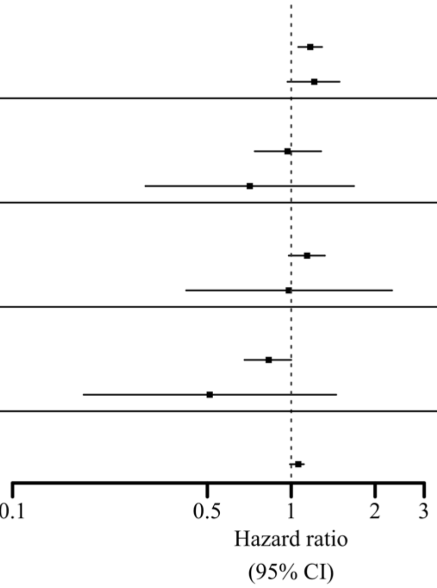


CI indicates confidence interval.
No bootstrapping method applied for the confidence interval.
[†] Derived using unweighted genetic risk score for each sleep trait, with adjustment for age, gender, assessment centre, 40 genetic principal components, and genotyping chip.
[‡] Derived using weighted genetic risk score for each sleep trait, with adjustment for age, gender, 20 genetic principal components, and genotyping chip.
[#] Hazard ratio (95% CI) scaled to per doubling in odds of the sleep trait. Chronotype was missing in HUNT2.

*Figure S11: 2x2 factorial Mendelian randomization Cox regression analysis assessing the joint effects of two sleep traits with risk of incident acute myocardial infarction in UK Biobank and HUNT2 after excluding participants who reported self-reported use of sleep medication.*

| | UK Biobank[†] | | HUNT2[†] | |
|---|---|---|---|---|
| | **N (incident cases)** | **Hazard ratio (95% CI)*** | **N (incident cases)** | **Hazard ratio (95% CI)*** |
| **Insomnia symptoms – Short sleep** | | | | |
| Both GRS ≤ median | 81 251 (1 769) | 1 [Reference] | 8 529 (764) | 1 [Reference] |
| Insomnia GRS > median | 70 993 (1 577) | 1.03 (0.96, 1.10) | 7 309 (653) | 1.05 (0.95, 1.17) |
| Short sleep GRS > median | 71 040 (1 637) | 1.05 (0.98, 1.12) | 7 341 (637) | 1.00 (0.90, 1.12) |
| Both GRS > median | 81 061 (1 950) | 1.10 (1.04, 1.18) | 8 430 (743) | 1.02 (0.92, 1.13) |
| **Insomnia symptoms – Long sleep** | | | | |
| Both GRS ≤ median | 72 818 (1 644) | 1 [Reference] | 9 606 (914) | 1 [Reference] |
| Insomnia GRS > median | 74 393 (1 788) | 1.07 (1.00, 1.15) | 9 957 (988) | 1.04 (0.95, 1.14) |
| Long sleep GRS > median | 53 186 (1 154) | 0.97 (0.90, 1.05) | 10 076 (940) | 0.94 (0.86, 1.03) |
| Both GRS > median | 51 463 (1 121) | 0.97 (0.90, 1.04) | 9 528 (906) | 1.02 (0.93, 1.12) |
| **Insomnia symptoms – Chronotype[#]** | | | | |
| Both GRS ≤ median | 79 595 (1 803) | 1 [Reference] | | |
| Insomnia GRS > median | 85 133 (1 963) | 1.03 (0.97, 1.10) | | |
| Chronotype GRS > median | 85 269 (1 978) | 1.03 (0.96, 1.10) | | |
| Both GRS > median | 79 437 (1 962) | 1.09 (1.03, 1.17) | | |
| **Short sleep – Chronotype[#]** | | | | |
| Both GRS ≤ median | 76 862 (1 667) | 1 [Reference] | | |
| Short sleep GRS > median | 75 326 (1 717) | 1.04 (0.97, 1.12) | | |
| Chronotype GRS > median | 75 382 (1 679) | 1.02 (0.96, 1.09) | | |
| Both GRS > median | 76 775 (1 870) | 1.11 (1.04, 1.19) | | |
| **Long sleep – Chronotype[#]** | | | | |
| Both GRS ≤ median | 71 549 (1 644) | 1 [Reference] | | |
| Long sleep GRS > median | 54 380 (1 187) | 0.95 (0.88, 1.03) | | |
| Chronotype GRS > median | 75 662 (1 788) | 1.03 (0.96, 1.10) | | |
| Both GRS > median | 50 269 (1 088) | 0.94 (0.87, 1.02) | | |

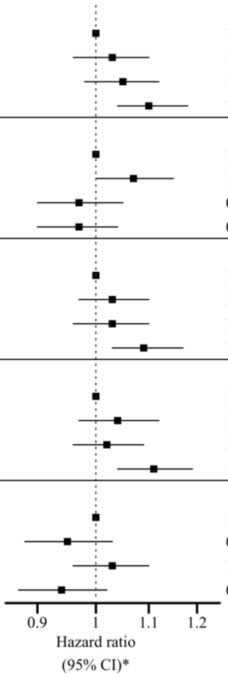


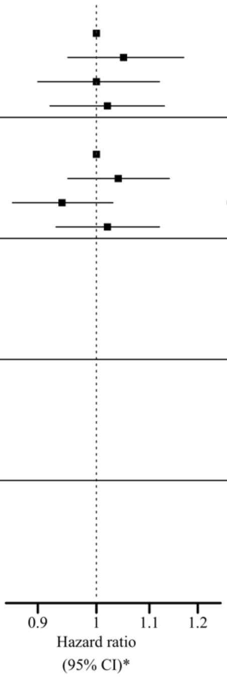


CI indicates confidence interval; and GRS, genetic risk score.

For each sleep trait combination, both GRS ≤ median represents low genetic risk for both sleep traits in combination, sleep trait 1 GRS > median represents high genetic risk for sleep trait 1 only, sleep trait 2 GRS > median represents high genetic risk for sleep trait 2 only and both GRS > median represents high genetic risk for both sleep traits.

[†] Derived using unweighted genetic risk score for each sleep trait in UK Biobank, whereas using weighted genetic risk score for each sleep trait in HUNT2.

[*] Adjusted for age, gender, assessment centre (in UK Biobank), genetic principal components (40 in UK Biobank and 20 in HUNT2), and genotyping chip.

[#] Chronotype genetic risk score calculated using alleles for morning preference.

#### Supplementary tables

*Table S1: Detailed summary of Mendelian randomization (MR) studies previously conducted on sleep traits and risk of coronary artery disease (CAD) or acute myocardial infarction (AMI).*

| Outcomes | Study | Design | Exposures | Results | Summary |
|---|---|---|---|---|---|
| **Coronary artery disease (CAD)*** | Daghlas et al. [8] | **Two-sample MR:**<br>Sleep duration SNPs from GWAS by Dashti et al. [14], and CAD summary GWAS data from the CARDIoGRAMplusC4D Consortium (60 801 cases and 123 504 controls).<br><br>**One-sample MR:**<br>Unweighted genetic instruments derived from sleep duration SNPs from Dashti et al. [14], and CAD from UK Biobank (17 157 cases and 320 375 controls) | Sleep duration and short sleep.<br><br>(Did not test long sleep in MR given the limited number of SNPs). | **Two-sample MR**<br>Sleep duration (per additional hour of sleep) IVW OR 0.79 (95% CI 0.68, 0.92); and<br><br>Short sleep IVW OR 1.24 (95% CI 1.11, 1.38)<br><br>**One-sample MR**<br>Sleep duration (per additional hour of sleep) OR 0.81 (95% CI 0.68, 0.97); and<br><br>Short sleep OR 1.14 (95% CI 1.03, 1.26) | Genetically predicted per additional hour of sleep was associated with significantly lower odds of CAD.<br><br>Short sleep a causal risk factor for CAD. |
| | Ai et al. [17] | **One-sample MR:**<br>Unweighted genetic risk scores derived from sleep duration SNPs from Dashti et al. [14], and CAD from UK Biobank (17 655 cases/404 044 UK Biobank samples) | Sleep duration, short sleep and long sleep. | Sleep duration (per additional hour of sleep) TSPS OR 0.80 (95% CI 0.66, 0.97);<br><br>Short sleep (15 594 cases) TSPS OR 1.24 (95% CI 1.12, 1.37); and<br><br>Long sleep (12 788 cases) TSPS OR 0.88 (95% CI 0.69, 1.34) | Genetically predicted per additional hour of sleep was associated with a lower odds of CAD.<br><br>Short sleep was associated with significantly higher odds of CAD. |
| | Larsson et al. [18] | **Two-sample MR:**<br>Insomnia SNPs from GWAS by Jansen et al. [13], and CAD summary GWAS data from the CARDIoGRAMplusC4D Consortium (n = 184 305 individuals of primarily (77%) European ancestry)) [19]. | Insomnia | IVW OR 1.12 (95% CI 1.08, 1.17) | Genetically predicted insomnia was associated with significantly higher odds of CAD. |
| | Liu et al. [20] | **Two-sample MR:**<br>Insomnia SNPs from GWAS by Jansen et al. [13], and CAD summary data derived based in imputed genotype data from the UK Biobank (32 463 cases/278 757 UK Biobank samples).<br><br>Replication using CAD summary GWAS data from the CARDIoGRAMplusC4D Consortium [19]. | Insomnia | IVW OR 1.22 (95% CI 1.17, 1.27); and<br><br>Replication IVW OR 1.13 (95% CI 1.08, 1.18) | Genetically predicted insomnia was significantly positively associated with CAD. |

| | | | | | |
|---|---|---|---|---|---|
| | Yuan et al. [21] | **Two-sample MR:**<br>Insomnia SNPs from GWAS by Jansen et al. [13], and CAD summary data derived based in imputed genotype data from the UK Biobank (29 278 cases and 338 308 controls). | Insomnia | IVW OR 1.19 (95% CI 1.14, 1.25) | Genetic liability to insomnia was associated with higher odds of CAD. |
| | Lane et al. [16] | **Two-sample MR:**<br>Insomnia SNPs from GWAS by Lane et al. [16], and CAD summary GWAS data from the CARDIoGRAMplusC4D Consortium [19].<br><br>**One-sample MR:**<br>Insomnia SNPs from GWAS by Lane et al. [16], and CAD summary data derived based in imputed genotype data from the UK Biobank (23 980 cases and 361 706 controls). | Insomnia<br>(any insomnia symptoms i.e., “sometimes”/“usually” as cases versus “never/rarely” as controls) | **Two-sample MR**<br>IVW OR 2.15 (95% CI 1.38, 3.35)<br><br>**One-sample MR**<br>OR 2.95 (95% CI 2.18, 3.99) | Genetically predicted insomnia was significantly positively associated with CAD. |
| **Acute myocardial infarction (AMI)** | Daghlas et al. [8] | **Two-sample MR:**<br>Sleep duration SNPs from GWAS by Dashti et al. [14], and AMI summary GWAS data from the CARDIoGRAMplusC4D Consortium with no participant overlap with UK Biobank (43 878 cases and 128 199 controls).<br><br>**One-sample MR:**<br>Weighted genetic instruments derived from sleep duration SNPs from Dashti et al. [14], and AMI from UK Biobank (12 111 cases and 325 421 controls) | Sleep duration and short sleep.<br><br>(Did not test long sleep in MR given the limited number of SNPs). | **Two-sample MR**<br>Sleep duration (per additional hour of sleep) IVW OR 0.80 (95% CI 0.67, 0.95)<br><br>Short sleep IVW OR 1.19 (95% CI 1.09, 1.29)<br><br>**One-sample MR**<br>Sleep duration (per additional hour of sleep) OR 0.86 (95% CI 0.70, 1.06)<br><br>Short sleep OR 1.21 (95% CI 1.08, 1.37) | Genetically predicted per additional hour of sleep was associated with lower odds of AMI.<br><br>Short sleep a causal risk factor for AMI. |
| | Ai et al. [17] | **One-sample MR:**<br>Unweighted genetic risk scores derived from sleep duration SNPs from Dashti et al.[14], and AMI from UK Biobank (16 845 cases/404 044 UK Biobank samples) | Sleep duration, short sleep and long sleep. | Sleep duration (per additional hour of sleep) TSPS OR 0.90 (95% CI 0.74, 1.09);<br><br>Short sleep (14 871 cases) TSPS OR 1.21 (95% CI 1.09, 1.34); and<br><br>Long sleep (12 206 cases) TSPS OR 0.94 (0.73, 1.22) | A weak evidence of protective effect of sleep duration on AMI.<br><br>Short sleep was associated with significantly higher odds of AMI. |
| | Yang et al. [22] | **Two-sample MR:**<br>Insomnia SNPs from Lane et al. [16], sleep duration SNPs from Dashti et al. [14], chronotype SNPs (111 SNPs only significant in UK Biobank) from Jones et al. [15]; and GWAS data for AMI | Insomnia, sleep duration, short sleep, long sleep and chronotype | Insomnia IVW OR 1.0049 (95% CI 1.0019, 1.0079);<br><br>Sleep duration (per additional hour of sleep) IVW OR 0.9999 (95% CI 0.9998, 1.0000); | Genetically predicted insomnia was significantly positively associated with AMI. |

| | | | | | |
|---|---|---|---|---|---|
| | | released by UK Biobank (7 018 cases and 354 176 controls). | | Short sleep IVW OR 1.0040 (95% CI 0.9989, 1.0091);<br><br>Long sleep IVW OR 0.9971 (95% CI 0.9910, 1.0031); and<br><br>Chronotype IVW OR 0.9992 (95% CI 0.9957, 1.0026). | A suggestive evidence that genetically predicted per additional hour of sleep was negatively associated with AMI. |

SNPs indicates single nucleotide polymorphisms; TSPS, two-stage predictor substitution; IVW, inverse variance weighted; OR, odds ratio; CI, confidence interval; GWAS, genome-wide association study; CAD, coronary artery disease; and AMI, acute myocardial infarction.

* CAD diagnosis was broadly defined as myocardial infarction, acute coronary syndrome, chronic stable angina or coronary stenosis of more than 50% [19].

*Table S2: Summary of genetic instruments showing their strength applying to UK Biobank and HUNT2.*

**UK Biobank**

| **Sleep traits** | **N** | **No. of SNPs to generate the uwGRS** | **Mean (SD) no. of increasing allele** | **Association of uwGRS with sleep trait †** | | |
|---|---|---|---|---|---|---|
| | | | | **Coefficient (SE)** | **$R^2$ §** | **F-statistics §§** |
| Insomnia symptoms | 332 676 | 248 | 245.17 (10.27) | 0.1560 (0.0039) | 0.41% | 1370.92 |
| 24-hour sleep duration (h) | 332 676 | 78 | 76.26 (5.43) | 0.0825 (0.0019) | 0.59% | 1962.0 |
| Short sleep (≤6 h vs. 7-8 h) | 307 135 | 27 | 26.34 (3.15) | 0.1044 (0.0041) | 0.18% | 558.68 |
| Long sleep (≥9 h vs. 7-8 h) | 253 811 | 8 | 4.11 (1.42) | 0.0900 (0.0066) | 0.11% | 285.42 |
| Chronotype (morning preference) | 332 676 | 341 # | 334.12 (11.64) | 0.2998 (0.0037) | 1.54% | 5202.20 |

**HUNT2**

| **Sleep traits** | **N** | **No. of SNPs to generate the wGRS** | **Mean (SD) no. of increasing allele** | **Association of wGRS with sleep trait ‡** | | |
|---|---|---|---|---|---|---|
| | | | | **Coefficient (SE)** | **$R^2$ §** | **F-statistics §§** |
| Insomnia symptoms | 44 728 | 244 # | 240.41 (10.16) | 0.1036 (0.0137) | 0.16% | 71.17 |
| 24-hour sleep duration (h) | 44 728 | 78 | 77.25 (5.35) | 0.0361 (0.0058) | 0.09% | 38.94 |
| Short sleep (≤6 h vs. 7-8 h) | 33 243 | 27 | 26.12 (3.13) | 0.0335 (0.0198) | 0.01% | 4.97 |
| Long sleep (≥9 h vs. 7-8 h) | 41 945 | 8 | 4.18 (1.40) | 0.0239 (0.0109) | 0.01% | 4.07 |

SNPs indicates single nucleotide polymorphisms; SD, standard deviation; uwGRS, unweighted genetic risk score; wGRS, weighted genetic risk score; and SE, standard error

† Adjusted for age, gender, assessment centre, 40 principal components, and genotyping chip.

‡ Adjusted for age, gender, 20 principal components, and genotyping chip.

§ McFadden $R^2$ statistics for sleep traits – insomnia symptoms, short sleep, long sleep and chronotype.

§§ F-statistics was calculated using $F = (R^2/K) / ((1 - R^2) / (N-K-1))$; where $R^2$ = McFadden $R^2$ statistics, K = 1, and N = sample size.

### rs146820337, rs112201801, rs10610420, rs9991917, rs67169439, rs34125199, rs60521023, rs3747463, rs213462, and rs7060620 were absent in the imputed UK Biobank genetic data; and rs1264419, rs138678612, rs238869 and rs3131638 were absent in the imputed HUNT genetic data.

*Table S3: Baseline characteristics of participants across groups categorized by dichotomizing to the median genetic risk scores for insomnia symptoms and short sleep in UK Biobank.*

| | UK Biobank (N = 307 135) | | | |
|---|---|---|---|---|
| | **Both GRS ≤ median** | **Insomnia GRS > median** | **Short sleep GRS > median** | **Both GRS > median** |
| **Total, % (n)** | 26.66 (81 895) | 23.34 (71 674) | 23.34 (71 673) | 26.66 (81 893) |
| **Variables, % (n)** | | | | |
| Male | 44.78 (36 673) | 44.51 (31 900) | 44.91 (32 187) | 45.04 (36 885) |
| Missing, % (n) | - | - | - | - |
| Married | 74.53 (61 035) | 74.04 (53 069) | 74.36 (53 294) | 73.92 (60 538) |
| Missing, % (n) | 0.45 (368) | 0.50 (361) | 0.45 (322) | 0.50 (410) |
| Weekly alcohol intake | 51.43 (42 121) | 50.70 (36 341) | 51.25 (36 735) | 50.53 (41 379) |
| Missing, % (n) | 0.04 (35) | 0.06 (42) | 0.04 (26) | 0.05 (43) |
| Current smokers | 9.46 (7 751) | 10.40 (7 456) | 9.59 (6 876) | 10.67 (8 737) |
| Missing, % (n) | 0.28 (232) | 0.28 (200) | 0.31 (219) | 0.30 (248) |
| Highly physically active | 33.98 (27 824) | 33.39 (23 934) | 33.68 (24 140) | 33.41 (27 363) |
| Missing, % (n) | 17.20 (14 085) | 17.75 (12 723) | 17.21 (12 336) | 18.09 (14 814) |
| Tertiary education | 45.19 (37 005) | 43.28 (31 020) | 43.84 (31 420) | 42.34 (34 671) |
| Missing, % (n) | 0.69 (568) | 0.75 (541) | 0.71 (508) | 0.77 (631) |
| Shift workers | 5.15 (4 214) | 5.35 (3 831) | 5.26 (3 770) | 5.42 (4 442) |
| Missing, % (n) | 0.25 (208) | 0.24 (171) | 0.28 (204) | 0.29 (236) |
| Employed | 59.37 (48 625) | 59.00 (42 286) | 58.71 (42 082) | 58.65 (48 034) |
| Missing, % (n) | 0.22 (184) | 0.21 (153) | 0.24 (175) | 0.24 (200) |
| Use of sleep medication(s) | 0.79 (644) | 0.95 (681) | 0.88 (633) | 1.02 (832) |
| Missing, % (n) | - | - | - | - |
| Suffering from depression | 10.56 (8 648) | 12.14 (8 704) | 10.82 (7 756) | 12.34 (10 107) |
| Missing, % (n) | - | - | - | - |
| Suffering from anxiety | 6.12 (5 016) | 6.79 (4 866) | 6.23 (4 463) | 6.80 (5 572) |
| Missing, % (n) | - | - | - | - |
| Suffering from chronic illness | 28.48 (23 325) | 31.18 (22 346) | 29.06 (20 831) | 31.98 (26 189) |
| Missing, % (n) | 1.92 (1 571) | 2.13 (1 528) | 1.95 (1 399) | 2.15 (1 759) |
| **Variables, mean (SD)** | | | | |
| Age, *years* | 56.75 (7.92) | 56.68 (7.94) | 56.83 (7.90) | 56.69 (7.95) |
| Missing, % (n) | - | - | - | - |
| TDI | -1.67 (2.87) | -1.57 (2.92) | -1.65 (2.89) | -1.54 (2.94) |
| Missing, % (n) | 0.11 (92) | 0.10 (74) | 0.13 (93) | 0.13 (104) |
| BMI, $kg/m^2$ | 27.16 (4.63) | 27.43 (4.77) | 27.24 (4.67) | 27.53 (4.81) |
| Missing, % (n) | 0.27 (220) | 0.31 (219) | 0.31 (220) | 0.30 (245) |
| SBP, *mmHg* | 138.10 (18.65) | 138.30 (18.61) | 138.20 (18.55) | 138.40 (18.57) |
| Missing, % (n) | 0.08 (69) | 0.10 (72) | 0.08 (55) | 0.08 (65) |
| Serum cholesterol, *mmol/L* | 5.76 (1.13) | 5.74 (1.13) | 5.75 (1.13) | 5.72 (1.13) |
| Missing, % (n) | 4.54 (3 722) | 4.60 (3 299) | 4.58 (3 282) | 4.44 (3 635) |
| Blood glucose, *mmol/L* | 5.09 (1.12) | 5.11 (1.21) | 5.10 (1.15) | 5.11 (1.19) |
| Missing, % (n) | 12.70 (10 397) | 12.76 (9 144) | 12.73 (9 124) | 12.70 (10 400) |

GRS indicates genetic risk score; SD, standard deviation; TDI, Townsend deprivation index; BMI, body mass index; and SBP, systolic blood pressure.

*Table S4: Baseline characteristics of participants across groups categorized by dichotomizing to the median genetic risk scores for insomnia symptoms and short sleep in HUNT2.*

| | **HUNT2 (N = 33 243)** | | | |
|---|---|---|---|---|
| | **Both GRS ≤ median** | **Insomnia GRS > median** | **Short sleep GRS > median** | **Both GRS > median** |
| **Total, % (n)** | 26.85 (8 925) | 23.15 (7 697) | 23.15 (7 697) | 26.84 (8 924) |
| **Variables, % (n)** | | | | |
| Male | 48.07 (4 290) | 47.46 (3 653) | 47.65 (3 668) | 46.62 (4 160) |
| Missing, % (n) | - | - | - | - |
| Married | 64.17 (5 727) | 64.09 (4 933) | 62.69 (4 825) | 62.82 (5 606) |
| Missing, % (n) | - | - | - | - |
| Weekly alcohol intake | 23.70 (2 115) | 23.97 (1 845) | 23.48 (1 807) | 22.98 (2 051) |
| Missing, % (n) | 6.89 (615) | 7.03 (541) | 7.03 (541) | 7.33 (654) |
| Current smokers | 27.59 (2 462) | 29.96 (2 306) | 27.22 (2 095) | 29.71 (2 651) |
| Missing, % (n) | 1.30 (116) | 1.51 (116) | 1.33 (102) | 1.42 (127) |
| Highly physically active | 35.87 (3 201) | 34.42 (2 649) | 35.07 (2 699) | 33.54 (2 993) |
| Missing, % (n) | 6.42 (573) | 6.31 (486) | 6.66 (513) | 7.26 (648) |
| Tertiary education | 23.10 (2 062) | 22.96 (1 767) | 22.35 (1 720) | 21.91 (1 955) |
| Missing, % (n) | 2.78 (248) | 2.64 (203) | 2.96 (228) | 3.08 (275) |
| Shift workers | 16.01 (1 429) | 17.02 (1 310) | 16.72 (1 287) | 16.62 (1 483) |
| Missing, % (n) | 7.70 (687) | 7.73 (595) | 7.96 (613) | 7.66 (684) |
| Employed | 74.26 (6 628) | 74.54 (5 737) | 73.43 (5 652) | 72.93 (6 508) |
| Missing, % (n) | 0.86 (77) | 0.95 (73) | 0.77 (59) | 0.92 (82) |
| Use of sleep medication(s) | 4.44 (396) | 5.04 (388) | 4.63 (356) | 5.54 (494) |
| Missing, % (n) | 9.30 (830) | 9.37 (721) | 9.80 (754) | 9.04 (807) |
| Suffering from chronic illness | 27.51 (2 455) | 28.18 (2 169) | 26.21 (2 017) | 29.66 (2 657) |
| Missing, % (n) | 2.80 (250) | 2.86 (220) | 2.91 (224) | 3.09 (276) |
| **Variables, mean (SD)** | | | | |
| Age, *years* | 47.49 (14.94) | 46.97 (14.81) | 47.04 (15.06) | 47.26 (15.04) |
| Missing, % (n) | - | - | - | - |
| BMI, *kg/m²* | 26.12 (3.92) | 26.23 (3.91) | 26.14 (3.95) | 26.31 (4.08) |
| Missing, % (n) | 0.19 (17) | 0.30 (23) | 0.29 (22) | 0.28 (25) |
| SBP, *mmHg* | 135.40 (20.17) | 135.00 (19.94) | 135.50 (19.88) | 135.20 (20.08) |
| Missing, % (n) | 0.06 (5) | 0.09 (7) | 0.16 (12) | 0.12 (11) |
| Serum cholesterol, *mmol/L* | 5.82 (1.21) | 5.81 (1.24) | 5.80 (1.21) | 5.79 (1.22) |
| Missing, % (n) | 0.04 (4) | 0.16 (12) | 0.16 (12) | 0.10 (9) |
| Blood glucose, *mmol/L* | 5.33 (1.24) | 5.36 (1.28) | 5.34 (1.31) | 5.37 (1.32) |
| Missing, % (n) | 0.08 (7) | 0.18 (14) | 0.17 (13) | 0.16 (14) |
| HADS - D scores | 3.25 (2.87) | 3.27 (2.94) | 3.19 (2.88) | 3.28 (2.94) |
| Missing, % (n) | 6.04 (539) | 5.43 (418) | 6.09 (469) | 6.36 (568) |
| HADS - A scores | 4.12 (3.19) | 4.20 (3.27) | 4.03 (3.11) | 4.21 (3.26) |
| Missing, % (n) | 12.09 (1 079) | 12.08 (930) | 12.08 (930) | 12.72 (1 135) |

GRS indicates genetic risk score; SD, standard deviation; BMI, body mass index; SBP, systolic blood pressure; HADS – D scores, Hospital Anxiety and Depression Scale – Depression scores; and HADS – A scores, Hospital Anxiety and Depression Scale – Anxiety scores.

*Table S5: Baseline characteristics of participants across groups categorized by dichotomizing to the median genetic risk scores for insomnia symptoms and long sleep in UK Biobank.*

| | **UK Biobank (N = 253 811)** | | | |
|---|---|---|---|---|
| | **Both GRS ≤ median** | **Insomnia GRS > median** | **Long sleep GRS > median** | **Both GRS > median** |
| **Total, % (n)** | 28.88 (73 306) | 29.55 (75 001) | 21.12 (53 600) | 20.45 (51 904) |
| **Variables, % (n)** | | | | |
| Male | 44.60 (32 692) | 44.62 (33 468) | 43.84 (23 497) | 44.37 (23 030) |
| Missing, % (n) | - | - | - | - |
| Married | 76.11 (55 790) | 75.70 (56 777) | 75.70 (40 573) | 75.57 (39 226) |
| Missing, % (n) | 0.42 (305) | 0.43 (321) | 0.39 (208) | 0.47 (245) |
| Weekly alcohol intake | 51.53 (37 772) | 50.84 (38 131) | 51.26 (27 477) | 50.80 (26 367) |
| Missing, % (n) | 0.03 (24) | 0.04 (30) | 0.04 (23) | 0.06 (32) |
| Current smokers | 8.89 (6 520) | 9.87 (7 399) | 9.22 (4 940) | 9.96 (5 169) |
| Missing, % (n) | 0.30 (219) | 0.27 (206) | 0.26 (138) | 0.25 (129) |
| Highly physically active | 33.80 (24 774) | 33.48 (25 113) | 33.59 (18 004) | 32.96 (17 105) |
| Missing, % (n) | 16.78 (12 300) | 17.37 (13 026) | 16.92 (9070) | 17.59 (9 130) |
| Tertiary education | 45.22 (33 151) | 43.68 (32 759) | 44.90 (24 065) | 43.43 (22 540) |
| Missing, % (n) | 0.67 (494) | 0.75 (559) | 0.64 (342) | 0.69 (359) |
| Shift workers | 4.52 (3 315) | 4.58 (3 432) | 4.36 (2 338) | 4.53 (2 351) |
| Missing, % (n) | 0.28 (205) | 0.26 (193) | 0.26 (140) | 0.26 (134) |
| Employed | 56.18 (41 184) | 56.23 (42 174) | 56.08 (30 058) | 56.01 (29 070) |
| Missing, % (n) | 0.24 (177) | 0.22 (167) | 0.22 (120) | 0.22 (115) |
| Use of sleep medication(s) | 0.67 (488) | 0.81 (608) | 0.77 (414) | 0.85 (435) |
| Missing, % (n) | - | - | - | - |
| Suffering from depression | 10.90 (7 993) | 12.22 (9 163) | 10.64 (5 702) | 12.04 (6 248) |
| Missing, % (n) | - | - | - | - |
| Suffering from anxiety | 6.00 (4 402) | 6.60 (4 949) | 6.12 (3 282) | 6.71 (3 482) |
| Missing, % (n) | - | - | - | - |
| Suffering from chronic illness | 28.43 (20 843) | 31.19 (23 390) | 28.29 (15 166) | 31.00 (16 092) |
| Missing, % (n) | 1.74 (1 278) | 1.92 (1 443) | 1.88 (1 007) | 2.04 (1 059) |
| **Variables, mean (SD)** | | | | |
| Age, *years* | 57.00 (7.97) | 56.88 (8.02) | 56.96 (8.02) | 56.87 (8.05) |
| Missing, % (n) | - | - | - | - |
| TDI | -1.74 (2.83) | -1.66 (2.85) | -1.74 (2.84) | -1.66 (2.88) |
| Missing, % (n) | 0.13 (97) | 0.10 (78) | 0.10 (56) | 0.13 (69) |
| BMI, *$kg/m^2$* | 27.17 (4.62) | 27.44 (4.76) | 27.01 (4.50) | 27.27 (4.65) |
| Missing, % (n) | 0.28 (207) | 0.31 (232) | 0.31 (168) | 0.26 (137) |
| SBP, *mmHg* | 138.30 (18.69) | 138.50 (18.73) | 138.20 (18.78) | 138.50 (18.73) |
| Missing, % (n) | 0.07 (50) | 0.09 (69) | 0.09 (49) | 0.09 (47) |
| Serum cholesterol, *mmol/L* | 5.75 (1.14) | 5.72 (1.14) | 5.76 (1.14) | 5.73 (1.14) |
| Missing, % (n) | 4.54 (3 330) | 4.53 (3 394) | 4.67 (2 504) | 4.60 (2 387) |
| Blood glucose, *mmol/L* | 5.10 (1.17) | 5.12 (1.20) | 5.09 (1.13) | 5.11 (1.18) |
| Missing, % (n) | 12.61 (9 245) | 12.64 (9 483) | 12.96 (6 947) | 12.89 (6 692) |

GRS indicates genetic risk score; SD, standard deviation; TDI, Townsend deprivation index; BMI, body mass index; and SBP, systolic blood pressure.

*Table S6: Baseline characteristics of participants across groups categorized by dichotomizing to the median genetic risk scores for insomnia symptoms and long sleep in HUNT2.*

| | HUNT2 (N = 41 945) | | | |
|---|---|---|---|---|
| | **Both GRS ≤ median** | **Insomnia GRS > median** | **Long sleep GRS > median** | **Both GRS > median** |
| **Total, % (n)** | 24.42 (10 242) | 25.58 (10 731) | 25.58 (10 731) | 24.42 (10 241) |
| **Variables, % (n)** | | | | |
| Male | 45.19 (4 628) | 45.01 (4 830) | 45.34 (4 865) | 44.26 (4 533) |
| Missing, % (n) | - | - | - | - |
| Married | 62.93 (6 445) | 63.42 (6 806) | 63.03 (6 764) | 61.83 (6 332) |
| Missing, % (n) | - | - | - | - |
| Weekly alcohol intake | 22.13 (2 267) | 21.98 (2 359) | 22.24 (2 387) | 22.46 (2 300) |
| Missing, % (n) | 7.21 (738) | 7.48 (803) | 7.64 (820) | 7.79 (798) |
| Current smokers | 26.29 (2 693) | 27.90 (2 994) | 26.14 (2 805) | 28.85 (2 955) |
| Missing, % (n) | 1.59 (163) | 1.53 (164) | 1.61 (173) | 1.50 (154) |
| Highly physically active | 33.26 (3 407) | 31.55 (3 386) | 32.36 (3 473) | 31.83 (3 260) |
| Missing, % (n) | 7.90 (809) | 8.09 (868) | 8.12 (871) | 8.25 (845) |
| Tertiary education | 20.92 (2 143) | 20.63 (2 214) | 21.59 (2 317) | 21.18 (2 169) |
| Missing, % (n) | 3.63 (372) | 3.41 (366) | 3.56 (382) | 3.65 (374) |
| Shift workers | 15.26 (1 563) | 14.84 (1 593) | 14.24 (1 528) | 15.39 (1 576) |
| Missing, % (n) | 7.08 (725) | 6.98 (749) | 7.43 (797) | 7.28 (746) |
| Employed | 66.71 (6 832) | 65.47 (7 026) | 65.35 (7 013) | 66.46 (6 806) |
| Missing, % (n) | 0.84 (86) | 0.94 (101) | 0.90 (97) | 0.98 (100) |
| Use of sleep medication(s) | 6.21 (636) | 7.21 (774) | 6.10 (655) | 6.96 (713) |
| Missing, % (n) | 9.78 (1 002) | 9.32 (1 000) | 9.69 (1 040) | 9.26 (948) |
| Suffering from chronic illness | 30.47 (3 121) | 33.35 (3 579) | 31.91 (3 424) | 32.55 (3 333) |
| Missing, % (n) | 3.12 (320) | 3.08 (330) | 3.04 (326) | 3.22 (330) |
| **Variables, mean (SD)** | | | | |
| Age, *years* | 49.17 (16.37) | 49.26 (16.27) | 49.57 (16.43) | 48.84 (16.33) |
| Missing, % (n) | - | - | - | - |
| BMI, *kg/m²* | 26.27 (4.06) | 26.39 (4.10) | 26.18 (3.98) | 26.32 (4.08) |
| Missing, % (n) | 0.51 (52) | 0.56 (60) | 0.51 (55) | 0.47 (48) |
| SBP, *mmHg* | 137.00 (21.24) | 136.70 (21.01) | 137.30 (21.27) | 136.70 (21.00) |
| Missing, % (n) | 0.09 (9) | 0.12 (13) | 0.12 (13) | 0.12 (12) |
| Serum cholesterol, *mmol/L* | 5.88 (1.25) | 5.88 (1.26) | 5.90 (1.25) | 5.86 (1.25) |
| Missing, % (n) | 0.12 (12) | 0.15 (16) | 0.09 (10) | 0.08 (8) |
| Blood glucose, *mmol/L* | 5.41 (1.46) | 5.44 (1.54) | 5.42 (1.41) | 5.41 (1.37) |
| Missing, % (n) | 0.17 (17) | 0.19 (20) | 0.13 (14) | 0.13 (13) |
| HADS - D scores | 3.31 (2.97) | 3.40 (3.01) | 3.34 (2.96) | 3.35 (2.96) |
| Missing, % (n) | 6.88 (705) | 6.64 (713) | 7.09 (761) | 6.76 (692) |
| HADS - A scores | 4.09 (3.18) | 4.18 (3.29) | 4.06 (3.18) | 4.18 (3.26) |
| Missing, % (n) | 13.70 (1 403) | 14.16 (1 519) | 13.91 (1 493) | 13.81 (1 414) |

GRS indicates genetic risk score; SD, standard deviation; BMI, body mass index; SBP, systolic blood pressure; HADS – D scores, Hospital Anxiety and Depression Scale – Depression scores; and HADS – A scores, Hospital Anxiety and Depression Scale – Anxiety scores.

*Table S7: Baseline characteristics of participants across groups categorized by dichotomizing to the median genetic risk scores for insomnia symptoms and chronotype (morning preference) in UK Biobank.*

| | **UK Biobank (N = 332 676)** | | | |
|---|---|---|---|---|
| | **Both GRS ≤ median** | **Insomnia GRS > median** | **Chronotype GRS > median** | **Both GRS > median** |
| **Total, % (n)** | 24.14 (80 295) | 25.86 (86 043) | 25.86 (86 043) | 24.14 (80 295) |
| **Variables, % (n)** | | | | |
| Male | 44.66 (35 857) | 44.39 (38 195) | 44.68 (38 440) | 44.87 (36 025) |
| Missing, % (n) | - | - | - | - |
| Married | 74.06 (59 465) | 73.55 (63 288) | 74.70 (64 270) | 74.29 (59 650) |
| Missing, % (n) | 0.47 (375) | 0.53 (459) | 0.48 (409) | 0.49 (391) |
| Weekly alcohol intake | 50.78 (40 777) | 49.92 (42 951) | 50.91 (43 804) | 50.30 (40 389) |
| Missing, % (n) | 0.04 (34) | 0.07 (60) | 0.05 (42) | 0.04 (31) |
| Current smokers | 9.74 (7 821) | 10.88 (9 363) | 9.55 (8 219) | 10.47 (8 409) |
| Missing, % (n) | 0.29 (236) | 0.29 (251) | 0.30 (260) | 0.30 (238) |
| Highly physically active | 33.14 (26 611) | 32.67 (28 110) | 33.98 (29 240) | 33.64 (27 010) |
| Missing, % (n) | 17.69 (14 206) | 18.17 (15 633) | 17.06 (14 676) | 17.99 (14 442) |
| Tertiary education | 44.21 (35 499) | 42.20 (36 309) | 43.77 (37 660) | 42.26 (33 936) |
| Missing, % (n) | 0.69 (553) | 0.74 (639) | 0.76 (650) | 0.83 (666) |
| Shift workers | 5.13 (4 119) | 5.36 (4 611) | 5.05 (4 343) | 5.15 (4 138) |
| Missing, % (n) | 0.28 (228) | 0.26 (227) | 0.27 (232) | 0.27 (218) |
| Employed | 57.10 (45 846) | 56.95 (49 003) | 56.96 (49 013) | 56.80 (45 606) |
| Missing, % (n) | 0.24 (194) | 0.22 (192) | 0.24 (207) | 0.24 (194) |
| Use of sleep medication(s) | 0.87 (700) | 1.06 (910) | 0.90 (774) | 1.07 (858) |
| Missing, % (n) | - | - | - | - |
| Suffering from depression | 11.38 (9 134) | 12.99 (11 180) | 11.34 (9 756) | 12.83 (10 300) |
| Missing, % (n) | - | - | - | - |
| Suffering from anxiety | 6.40 (5 138) | 7.02 (6 039) | 6.41 (5 516) | 7.02 (5 640) |
| Missing, % (n) | - | - | - | - |
| Suffering from chronic illness | 30.00 (24 086) | 32.94 (28 344) | 29.53 (25 406) | 32.42 (26 035) |
| Missing, % (n) | 1.98 (1 591) | 2.17 (1 865) | 1.90 (1 638) | 2.09 (1 678) |
| **Variables, mean (SD)** | | | | |
| Age, *years* | 56.93 (7.94) | 56.82 (7.97) | 56.98 (7.91) | 56.87 (7.94) |
| Missing, % (n) | - | - | - | - |
| TDI | -1.60 (2.92) | -1.51 (2.96) | -1.68 (2.87) | -1.56 (2.92) |
| Missing, % (n) | 0.10 (82) | 0.13 (108) | 0.13 (114) | 0.11 (91) |
| BMI, *kg/m²* | 27.23 (4.66) | 27.54 (4.83) | 27.27 (4.70) | 27.55 (4.83) |
| Missing, % (n) | 0.31 (246) | 0.30 (262) | 0.30 (257) | 0.32 (260) |
| SBP, *mmHg* | 138.30 (18.72) | 138.50 (18.73) | 138.30 (18.57) | 138.40 (18.53) |
| Missing, % (n) | 0.08 (68) | 0.10 (85) | 0.09 (76) | 0.09 (73) |
| Serum cholesterol, *mmol/L* | 5.75 (1.14) | 5.73 (1.14) | 5.75 (1.13) | 5.73 (1.13) |
| Missing, % (n) | 4.55 (3 651) | 4.48 (3 853) | 4.61 (3 966) | 4.62 (3 710) |
| Blood glucose, *mmol/L* | 5.11 (1.19) | 5.13 (1.22) | 5.10 (1.13) | 5.12 (1.23) |
| Missing, % (n) | 12.75 (10 236) | 12.58 (10 823) | 12.71 (10 934) | 12.87 (10 337) |

GRS indicates genetic risk score; SD, standard deviation; TDI, Townsend deprivation index; BMI, body mass index; and SBP, systolic blood pressure.

*Table S8: Baseline characteristics of participants across groups categorized by dichotomizing to the median genetic risk scores for short sleep and chronotype (morning preference) in UK Biobank.*

| | **UK Biobank (N = 307 135)** | | | |
|---|---|---|---|---|
| | **Both GRS ≤ median** | **Short sleep GRS > median** | **Chronotype GRS > median** | **Both GRS > median** |
| **Total, % (n)** | 25.24 (77 511) | 24.76 (76 057) | 24.76 (76 058) | 25.24 (77 509) |
| **Variables, % (n)** | | | | |
| Male | 44.52 (34 506) | 44.87 (34 128) | 44.79 (34 067) | 45.08 (34 944) |
| Missing, % (n) | - | - | - | - |
| Married | 74.07 (57 414) | 73.70 (56 055) | 74.54 (56 690) | 74.54 (57 777) |
| Missing, % (n) | 0.49 (376) | 0.48 (368) | 0.46 (353) | 0.47 (364) |
| Weekly alcohol intake | 50.96 (39 500) | 50.77 (38 611) | 51.23 (38 962) | 50.97 (39 503) |
| Missing, % (n) | 0.06 (48) | 0.05 (36) | 0.04 (29) | 0.04 (33) |
| Current smokers | 10.02 (7 766) | 10.37 (7 888) | 9.78 (7 441) | 9.97 (7 725) |
| Missing, % (n) | 0.28 (216) | 0.31 (235) | 0.28 (216) | 0.30 (232) |
| Highly physically active | 33.25 (25 770) | 33.14 (25 202) | 34.17 (25 988) | 33.93 (26 301) |
| Missing, % (n) | 17.70 (13 718) | 17.87 (13 592) | 17.21 (13 090) | 17.49 (13 558) |
| Tertiary education | 44.24 (34 289) | 43.23 (32 883) | 44.36 (33 736) | 42.84 (33 208) |
| Missing, % (n) | 0.68 (527) | 0.71 (543) | 0.77 (582) | 0.77 (596) |
| Shift workers | 5.28 (4 096) | 5.42 (4 123) | 5.19 (3 949) | 5.28 (4 089) |
| Missing, % (n) | 0.24 (183) | 0.30 (226) | 0.26 (196) | 0.28 (214) |
| Employed | 59.31 (45 968) | 58.80 (44 722) | 59.09 (44 943) | 58.57 (45 394) |
| Missing, % (n) | 0.20 (157) | 0.24 (186) | 0.24 (180) | 0.24 (189) |
| Use of sleep medication(s) | 0.84 (649) | 0.96 (731) | 0.89 (676) | 0.95 (734) |
| Missing, % (n) | - | - | - | - |
| Suffering from depression | 11.35 (8 797) | 11.76 (8 943) | 11.25 (8 555) | 11.51 (8 920) |
| Missing, % (n) | - | - | - | - |
| Suffering from anxiety | 6.50 (5 042) | 6.47 (4 919) | 6.36 (4 840) | 6.60 (5 116) |
| Missing, % (n) | - | - | - | - |
| Suffering from chronic illness | 30.02 (23 266) | 30.84 (23 453) | 29.46 (22 405) | 30.41 (23 567) |
| Missing, % (n) | 2.06 (1 600) | 2.09 (1 588) | 1.97 (1 499) | 2.03 (1 570) |
| **Variables, mean (SD)** | | | | |
| Age, *years* | 56.67 (7.96) | 56.74 (7.93) | 56.76 (7.91) | 56.77 (7.92) |
| Missing, % (n) | - | - | - | - |
| TDI | -1.59 (2.91) | -1.55 (2.94) | -1.66 (2.88) | -1.62 (2.89) |
| Missing, % (n) | 0.10 (81) | 0.12 (95) | 0.11 (85) | 0.13 (102) |
| BMI, *kg/m$^2$* | 27.30 (4.69) | 27.38 (4.74) | 27.28 (4.70) | 27.40 (4.75) |
| Missing, % (n) | 0.30 (232) | 0.29 (222) | 0.27 (207) | 0.31 (243) |
| SBP, *mmHg* | 138.20 (18.73) | 138.40 (18.66) | 138.20 (18.53) | 138.30 (18.47) |
| Missing, % (n) | 0.09 (71) | 0.08 (61) | 0.09 (70) | 0.08 (59) |
| Serum cholesterol, *mmol/L* | 5.75 (1.13) | 5.74 (1.14) | 5.75 (1.13) | 5.74 (1.13) |
| Missing, % (n) | 4.52 (3 500) | 4.49 (3 412) | 4.63 (3 521) | 4.52 (3 505) |
| Blood glucose, *mmol/L* | 5.10 (1.18) | 5.11 (1.18) | 5.10 (1.14) | 5.10 (1.17) |
| Missing, % (n) | 12.73 (9 865) | 12.62 (9 600) | 12.72 (9 676) | 12.80 (9 924) |

GRS indicates genetic risk score; SD, standard deviation; TDI, Townsend deprivation index; BMI, body mass index; and SBP, systolic blood pressure.

*Table S9: Baseline characteristics of participants across groups categorized by dichotomizing to the median genetic risk scores for long sleep and chronotype (morning preference) in UK Biobank.*

| | **UK Biobank (N = 253 811)** | | | |
|---|---|---|---|---|
| | **Both GRS ≤ median** | **Long sleep GRS > median** | **Chronotype GRS > median** | **Both GRS > median** |
| **Total, % (n)** | 28.40 (72 088) | 21.60 (54 818) | 30.03 (76 219) | 19.97 (50 686) |
| **Variables, % (n)** | | | | |
| Male | 44.52 (32 092) | 43.95 (24 093) | 44.70 (34 068) | 44.26 (22 434) |
| Missing, % (n) | - | - | - | - |
| Married | 75.49 (54 418) | 75.46 (41 363) | 76.29 (58 149) | 75.83 (38 436) |
| Missing, % (n) | 0.42 (305) | 0.45 (245) | 0.42 (321) | 0.41 (208) |
| Weekly alcohol intake | 51.00 (36 762) | 50.94 (27 923) | 51.35 (39 141) | 51.14 (25 921) |
| Missing, % (n) | 0.04 (29) | 0.06 (33) | 0.03 (25) | 0.04 (22) |
| Current smokers | 9.52 (6 862) | 9.75 (5 345) | 9.26 (7 057) | 9.40 (4 764) |
| Missing, % (n) | 0.28 (205) | 0.26 (143) | 0.29 (220) | 0.24 (124) |
| Highly physically active | 33.15 (23 896) | 33.01 (18 093) | 34.10 (25 991) | 33.57 (17 016) |
| Missing, % (n) | 17.30 (12 471) | 17.52 (9 605) | 16.87 (12 855) | 16.96 (8 595) |
| Tertiary education | 44.55 (32 116) | 44.14 (24 194) | 44.34 (33 794) | 44.22 (22 411) |
| Missing, % (n) | 0.70 (504) | 0.63 (347) | 0.72 (549) | 0.70 (354) |
| Shift workers | 4.65 (3 354) | 4.50 (2 466) | 4.45 (3 393) | 4.39 (2 223) |
| Missing, % (n) | 0.28 (199) | 0.27 (146) | 0.26 (199) | 0.25 (128) |
| Employed | 56.31 (40 594) | 55.96 (30 674) | 56.11 (42 764) | 56.14 (28 454) |
| Missing, % (n) | 0.23 (164) | 0.23 (125) | 0.24 (180) | 0.22 (110) |
| Use of sleep medication(s) | 0.75 (539) | 0.80 (438) | 0.73 (557) | 0.82 (417) |
| Missing, % (n) | - | - | - | - |
| Suffering from depression | 11.67 (8 411) | 11.42 (6 262) | 11.47 (8 745) | 11.22 (5 688) |
| Missing, % (n) | - | - | - | - |
| Suffering from anxiety | 6.42 (4 629) | 6.33 (3 472) | 6.20 (4 722) | 6.49 (3 292) |
| Missing, % (n) | - | - | - | - |
| Suffering from chronic illness | 30.21 (21 777) | 29.86 (16 369) | 29.46 (22 456) | 29.37 (14 889) |
| Missing, % (n) | 1.90 (1 369) | 2.03 (1 114) | 1.77 (1 352) | 1.88 (952) |
| **Variables, mean (SD)** | | | | |
| Age, *years* | 56.94 (8.00) | 56.90 (8.06) | 56.94 (8.00) | 56.92 (8.00) |
| Missing, % (n) | - | - | - | - |
| TDI | -1.67 (2.87) | -1.66 (2.88) | -1.73 (2.82) | -1.74 (2.83) |
| Missing, % (n) | 0.12 (90) | 0.10 (54) | 0.11 (85) | 0.14 (71) |
| BMI, *kg/m²* | 27.31 (4.69) | 27.13 (4.56) | 27.31 (4.70) | 27.15 (4.60) |
| Missing, % (n) | 0.28 (203) | 0.30 (162) | 0.31 (236) | 0.28 (143) |
| SBP, *mmHg* | 138.40 (18.84) | 138.40 (18.79) | 138.30 (18.60) | 138.20 (18.73) |
| Missing, % (n) | 0.09 (63) | 0.08 (46) | 0.07 (56) | 0.10 (50) |
| Serum cholesterol, *mmol/L* | 5.74 (1.14) | 5.75 (1.14) | 5.74 (1.14) | 5.74 (1.13) |
| Missing, % (n) | 4.48 (3 228) | 4.58 (2 508) | 4.59 (3 496) | 4.70 (2 383) |
| Blood glucose, *mmol/L* | 5.12 (1.20) | 5.12 (1.17) | 5.10 (1.17) | 5.09 (1.13) |
| Missing, % (n) | 12.51 (9 021) | 12.75 (6 990) | 12.74 (9 707) | 13.12 (6 649) |

GRS indicates genetic risk score; SD, standard deviation; TDI, Townsend deprivation index; BMI, body mass index; and SBP, systolic blood pressure.

*Table S10: Statistical test of the proportional hazard assumption for one-sample Mendelian randomization (MR) Cox regression models.*

| | UK Biobank † | | HUNT2 ‡ | |
|---|---|---|---|---|
| **Sleep trait** | **Correlation coefficient *** | **P** | **Correlation coefficient *** | **P** |
| Insomnia symptoms | -0.0047 | 0.676 | 0.0166 | 0.269 |
| 24-hour sleep duration (h) | 0.0067 | 0.558 | 0.0165 | 0.260 |
| Short sleep (≤6 h vs. 7-8 h) | -0.0070 | 0.560 | 0.0048 | 0.788 |
| Long sleep (≥9 h vs. 7-8 h) | -0.0081 | 0.539 | -0.0162 | 0.293 |
| Chronotype (morning preference) | 0.0033 | 0.772 | - | - |

† Using unweighted genetic risk score for each sleep trait in the MR Cox regression model, with adjustment for age, gender, assessment centre, 40 genetic principal components, and genotyping chip.

‡ Using weighted genetic risk score for each sleep trait in the MR Cox regression model, with adjustment for age, gender, 20 genetic principal components, and genotyping chip.

* Values represent the Pearson's correlation coefficient between the first scaled Schoenfeld residual in the MR Cox regression and the rank-normalised natural logarithm of follow-up time.

*Table S11: Statistical test of the proportional hazard assumption for 2x2 factorial Mendelian randomization (MR) Cox regression models.*

| | UK Biobank † | | HUNT2 ‡ | |
|---|---|---|---|---|
| **Sleep trait combination** | **Correlation coefficient *** | **P** | **Correlation coefficient *** | **P** |
| Insomnia symptoms – Short sleep | -0.0084 | 0.479 | 0.0259 | 0.153 |
| Insomnia symptoms – Long sleep | -0.0144 | 0.280 | -0.0016 | 0.917 |
| Insomnia symptoms – Chronotype # | 0.0029 | 0.795 | - | - |
| Short sleep – Chronotype # | -0.0061 | 0.610 | - | - |
| Long sleep – Chronotype # | 0.0051 | 0.701 | - | - |

† Using unweighted genetic risk score for the sleep traits in the factorial MR Cox regression model, with adjustment for age, gender, assessment centre, 40 genetic principal components, and genotyping chip.

‡ Using weighted genetic risk score for each sleep trait in the factorial MR Cox regression model, with adjustment for age, gender, 20 genetic principal components, and genotyping chip.

* Values represent the Pearson's correlation coefficient between the first scaled Schoenfeld residual in the factorial MR Cox regression and the rank-normalised natural logarithm of follow-up time.

### Chronotype genetic risk score calculated using alleles for morning preference.

*Table S12: One-sample Mendelian randomization Cox regression analysis for risk of incident acute myocardial infarction associated with sleep traits in HUNT2 using weighted and unweighted genetic risk scores for sleep traits.*

| | **Weighted genetic risk score** | | **Unweighted genetic risk score** | |
|---|---|---|---|---|
| **Sleep trait** | **N (incident cases)** | **Hazard ratio (95% CI)*** | **N (incident cases)** | **Hazard ratio (95% CI)*** |
| Insomnia symptoms # | 44 728 (4 488) | 1.23 (1.00, 1.55) | 44 728 (4 488) | 1.24 (1.00, 1.59) |
| 24-hour sleep duration (h) | 44 728 (4 488) | 0.76 (0.31, 1.79) | 44 728 (4 488) | 0.72 (0.30, 1.64) |
| Short sleep # (≤6 h vs. 7-8 h) | 33 243 (3 058) | 0.87 (0.15, 3.24) | 33 243 (3 058) | 0.82 (0.06, 6.53) |
| Long sleep # (≥9 h vs. 7-8 h) | 41 945 (4 209) | 0.53 (0.01, 8.28) | 41 945 (4 209) | 0.85 (0.29, 1.83) |

CI indicates confidence interval.

* Adjusted for age, gender, 20 genetic principal components, and genotyping chip.

### Hazard ratio (95% CI) scaled to per doubling in odds of the sleep trait.

*Table S13: Associations between genetic risk scores and potential confounders in UK Biobank.*

| Instrument | Confounder | Coefficient (Beta) | SE | N | P | |
|---|---|---|---|---|---|---|
| Insomnia uwGRS | Marital status | 0.057 | 0.051 | 230 590 | 0.270 | |
| | Alcohol intake | -0.083 | 0.023 | 230 590 | 3.34e-04 | * |
| | Smoking status | 0.286 | 0.033 | 230 590 | <2e-16 | * |
| | BMI | 0.053 | 0.005 | 230 590 | <2e-16 | * |
| | Physical activity | -0.014 | 0.029 | 230 590 | 0.625 | |
| | TDI | 0.030 | 0.008 | 230 590 | 1.46e-04 | * |
| | Education | -0.215 | 0.023 | 230 590 | <2e-16 | * |
| | Shift work | 0.061 | 0.102 | 230 590 | 0.547 | |
| | Employment status | 0.146 | 0.046 | 230 590 | 0.002 | |
| | SBP | 7.99e-04 | 0.001 | 230 590 | 0.512 | |
| | Fasting time | -0.002 | 0.009 | 230 590 | 0.786 | |
| | Serum cholesterol | -0.059 | 0.019 | 230 590 | 0.002 | |
| | Blood glucose | 0.020 | 0.018 | 230 590 | 0.280 | |
| | Depression | 0.632 | 0.071 | 230 590 | <2e-16 | * |
| | Anxiety | 0.279 | 0.092 | 230 590 | 0.002 | |
| | Sleep medication | 0.413 | 0.231 | 230 590 | 0.074 | |
| | Chronic illness | 0.683 | 0.049 | 230 590 | <2e-16 | * |
| Sleep duration uwGRS | Marital status | -0.009 | 0.027 | 230 590 | 0.742 | |
| | Alcohol intake | 0.001 | 0.012 | 230 590 | 0.910 | |
| | Smoking status | -0.014 | 0.017 | 230 590 | 0.410 | |
| | BMI | -0.016 | 0.003 | 230 590 | 1.62e-09 | * |
| | Physical activity | -0.009 | 0.016 | 230 590 | 0.553 | |
| | TDI | -0.004 | 0.004 | 230 590 | 0.307 | |
| | Education | 0.048 | 0.012 | 230 590 | 8.95e-05 | * |
| | Shift work | 0.004 | 0.054 | 230 590 | 0.939 | |
| | Employment status | -0.064 | 0.024 | 230 590 | 0.008 | |
| | SBP | 0.001 | 0.001 | 230 590 | 0.030 | |
| | Fasting time | 2.84e-04 | 0.005 | 230 590 | 0.953 | |
| | Serum cholesterol | -0.002 | 0.010 | 230 590 | 0.860 | |
| | Blood glucose | 0.002 | 0.010 | 230 590 | 0.842 | |
| | Depression | 0.064 | 0.038 | 230 590 | 0.089 | |
| | Anxiety | 0.030 | 0.049 | 230 590 | 0.535 | |
| | Sleep medication | -0.182 | 0.122 | 230 590 | 0.137 | |
| | Chronic illness | -0.065 | 0.026 | 230 590 | 0.013 | |
| Short sleep uwGRS | Marital status | 0.005 | 0.016 | 213 418 | 0.768 | |
| | Alcohol intake | -0.028 | 0.007 | 213 418 | 1.31e-04 | * |
| | Smoking status | 0.016 | 0.011 | 213 418 | 0.136 | |
| | BMI | 0.006 | 0.002 | 213 418 | 8.84e-05 | * |
| | Physical activity | -0.006 | 0.009 | 213 418 | 0.534 | |
| | TDI | 0.002 | 0.002 | 213 418 | 0.494 | |
| | Education | -0.060 | 0.007 | 213 418 | 1.38e-15 | * |
| | Shift work | -0.001 | 0.032 | 213 418 | 0.977 | |
| | Employment status | 0.004 | 0.015 | 213 418 | 0.760 | |
| | SBP | 4.93e-04 | 3.90e-04 | 213 418 | 0.206 | |
| | Fasting time | 0.001 | 0.003 | 213 418 | 0.732 | |
| | Serum cholesterol | -0.007 | 0.006 | 213 418 | 0.267 | |
| | Blood glucose | -0.006 | 0.006 | 213 418 | 0.352 | |
| | Depression | 0.042 | 0.023 | 213 418 | 0.068 | |
| | Anxiety | 0.002 | 0.029 | 213 418 | 0.837 | |
| | Sleep medication | 0.072 | 0.076 | 213 418 | 0.345 | |
| | Chronic illness | 0.040 | 0.016 | 213 418 | 0.011 | |
| Long sleep uwGRS | Marital status | -0.020 | 0.008 | 177 512 | 0.014 | |
| | Alcohol intake | 0.004 | 0.004 | 177 512 | 0.288 | |
| | Smoking status | 0.002 | 0.005 | 177 512 | 0.660 | |
| | BMI | -0.007 | 7.87e-04 | 177 512 | <2e-16 | * |
| | Physical activity | -0.019 | 0.005 | 177 512 | 4.58e-05 | * |
| | TDI | 4.06e-04 | 0.001 | 177 512 | 0.748 | |
| | Education | -0.005 | 0.004 | 177 512 | 0.153 | |
| | Shift work | -0.009 | 0.017 | 177 512 | 0.591 | |
| | Employment status | -0.004 | 0.007 | 177 512 | 0.558 | |
| | SBP | 2.27e-04 | 1.92e-04 | 177 512 | 0.236 | |
| | Fasting time | 0.002 | 0.001 | 177 512 | 0.141 | |
| | Serum cholesterol | 0.008 | 0.003 | 177 512 | 0.012 | |
| | Blood glucose | 3.81e-04 | 0.003 | 177 512 | 0.898 | |
| | Depression | -0.013 | 0.011 | 177 512 | 0.238 | |
| | Anxiety | 0.024 | 0.015 | 177 512 | 0.099 | |
| | Sleep medication | 0.030 | 0.040 | 177 512 | 0.453 | |
| | Chronic illness | -9.32e-05 | 0.008 | 177 512 | 0.991 | |
| Chronotype (morning preference) uwGRS | Marital status | 0.129 | 0.058 | 230 590 | 0.028 | |
| | Alcohol intake | -0.055 | 0.026 | 230 590 | 0.038 | |
| | Smoking status | -0.047 | 0.037 | 230 590 | 0.209 | |
| | BMI | 0.015 | 0.006 | 230 590 | 0.006 | |
| | Physical activity | 0.202 | 0.033 | 230 590 | 1.53e-09 | * |
| | TDI | -0.051 | 0.009 | 230 590 | 1.18e-08 | * |
| | Education | -0.070 | 0.026 | 230 590 | 0.008 | |
| | Shift work | -0.104 | 0.115 | 230 590 | 0.368 | |
| | Employment status | -0.027 | 0.052 | 230 590 | 0.601 | |
| | SBP | -0.001 | 0.001 | 230 590 | 0.331 | |

| | | | | | |
|---|---|---|---|---|---|
| Fasting time | -0.020 | 0.010 | 230 590 | 0.054 | |
| Serum cholesterol | -0.050 | 0.022 | 230 590 | 0.022 | |
| Blood glucose | -0.069 | 0.021 | 230 590 | 0.001 | |
| Depression | -0.002 | 0.080 | 230 590 | 0.980 | |
| Anxiety | 0.071 | 0.104 | 230 590 | 0.496 | |
| Sleep medication | -0.088 | 0.262 | 230 590 | 0.737 | |
| Chronic illness | -0.208 | 0.056 | 230 590 | 1.87e-04 | * |

uwGRS indicates unweighted genetic risk score; SE, standard error; BMI, body mass index; TDI, Townsend deprivation index; SBP, systolic blood pressure.
Coefficients are in terms of an average-SNP increase in the allele score per unit/level increase in confounder.
* Associations surpassing multiple-testing corrected p-value threshold of 0.05/85 = 5.88e-04.

*Table S14: Associations between genetic risk scores and potential confounders in HUNT2.*

| Instrument | Confounder | Coefficient (Beta) | SE | N | P | |
|---|---|---|---|---|---|---|
| Insomnia wGRS | Marital status | 0.004 | 0.005 | 27 309 | 0.364 | |
| | Alcohol intake | 0.001 | 0.004 | 27 309 | 0.757 | |
| | Smoking status | 0.015 | 0.003 | 27 309 | 9.47e-06 | * |
| | BMI | 0.003 | 0.001 | 27 309 | 7.07e-05 | * |
| | Physical activity | -0.007 | 0.003 | 27 309 | 0.029 | |
| | Education | -0.001 | 0.004 | 27 309 | 0.770 | |
| | Shift work | 0.008 | 0.007 | 27 309 | 0.272 | |
| | Employment status | 0.002 | 0.007 | 27 309 | 0.824 | |
| | SBP | -1.83e-04 | 1.61e-04 | 27 309 | 0.254 | |
| | Fasting time | 0.002 | 0.001 | 27 309 | 0.224 | |
| | Serum cholesterol | -0.009 | 0.002 | 27 309 | 2.02e-04 | * |
| | Blood glucose | -5.35e-04 | 0.002 | 27 309 | 0.805 | |
| | HADS - Depression | -0.004 | 0.001 | 27 309 | 0.003 | |
| | HADS - Anxiety | 0.004 | 0.001 | 27 309 | 2.58e-04 | * |
| | Sleep medication | 0.034 | 0.013 | 27 309 | 0.007 | |
| | Chronic illness | 0.013 | 0.007 | 27 309 | 0.058 | |
| Sleep duration wGRS | Marital status | -0.222 | 3.390 | 27 309 | 0.948 | |
| | Alcohol intake | 4.280 | 2.613 | 27 309 | 0.101 | |
| | Smoking status | -3.574 | 2.399 | 27 309 | 0.136 | |
| | BMI | -0.734 | 0.515 | 27 309 | 0.154 | |
| | Physical activity | 0.142 | 2.351 | 27 309 | 0.952 | |
| | Education | -3.158 | 2.971 | 27 309 | 0.288 | |
| | Shift work | -2.939 | 5.150 | 27 309 | 0.568 | |
| | Employment status | 2.412 | 4.930 | 27 309 | 0.625 | |
| | SBP | -0.040 | 0.113 | 27 309 | 0.721 | |
| | Fasting time | -0.830 | 1.023 | 27 309 | 0.417 | |
| | Serum cholesterol | -1.245 | 1.765 | 27 309 | 0.481 | |
| | Blood glucose | 0.727 | 1.520 | 27 309 | 0.632 | |
| | HADS - Depression | -0.330 | 0.838 | 27 309 | 0.694 | |
| | HADS - Anxiety | 0.077 | 0.740 | 27 309 | 0.917 | |
| | Sleep medication | 4.460 | 8.876 | 27 309 | 0.615 | |
| | Chronic illness | -2.337 | 4.692 | 27 309 | 0.618 | |
| Short sleep wGRS | Marital status | 4.61e-04 | 0.001 | 21 020 | 0.723 | |
| | Alcohol intake | -4.71e-04 | 0.001 | 21 020 | 0.638 | |
| | Smoking status | 0.001 | 9.15e-04 | 21 020 | 0.121 | |
| | BMI | 5.10e-04 | 2.01e-04 | 21 020 | 0.011 | |
| | Physical activity | -0.001 | 8.91e-04 | 21 020 | 0.249 | |
| | Education | -1.46e-04 | 0.001 | 21 020 | 0.898 | |
| | Shift work | -0.001 | 0.002 | 21 020 | 0.456 | |
| | Employment status | -0.003 | 0.002 | 21 020 | 0.115 | |
| | SBP | 2.11e-05 | 4.48e-05 | 21 020 | 0.638 | |
| | Fasting time | 4..73e-04 | 3.89e-04 | 21 020 | 0.224 | |
| | Serum cholesterol | -0.002 | 6.85e-04 | 21 020 | 0.021 | |
| | Blood glucose | 1.20e-04 | 6.40e-04 | 21 020 | 0.851 | |
| | HADS - Depression | -3.15e-04 | 3.28e-04 | 21 020 | 0.337 | |
| | HADS - Anxiety | -1.63e-04 | 2.86e-04 | 21 020 | 0.568 | |
| | Sleep medication | -7.08e-04 | 0.004 | 21 020 | 0.853 | |
| | Chronic illness | 8.66e-04 | 0.002 | 21 020 | 0.638 | |
| Long sleep wGRS | Marital status | -6.48e-04 | 0.001 | 25 635 | 0.535 | |
| | Alcohol intake | 5.24e-04 | 8.00e-04 | 25 635 | 0.512 | |
| | Smoking status | 2.82e-04 | 7.36e-04 | 25 635 | 0.702 | |
| | BMI | -3.87e-04 | 1.58e-04 | 25 635 | 0.014 | |
| | Physical activity | -3.64e-04 | 7.21e-04 | 25 635 | 0.614 | |
| | Education | 0.002 | 9.08e-04 | 25 635 | 0.052 | |
| | Shift work | 7.98e-05 | 0.002 | 25 635 | 0.960 | |
| | Employment status | -0.001 | 0.001 | 25 635 | 0.446 | |
| | SBP | 2.22e-05 | 3.45e-05 | 25 635 | 0.519 | |
| | Fasting time | -1.98e-04 | 3.15e-04 | 25 635 | 0.530 | |
| | Serum cholesterol | 2.42e-04 | 5.40e-04 | 25 635 | 0.654 | |
| | Blood glucose | -2.32e-04 | 4.63e-04 | 25 635 | 0.617 | |
| | HADS - Depression | 2.99e-04 | 2.59e-04 | 25 635 | 0.247 | |
| | HADS - Anxiety | -1.40e-04 | 2.28e-04 | 25 635 | 0.540 | |
| | Sleep medication | -0.003 | 0.003 | 25 635 | 0.231 | |
| | Chronic illness | 0.001 | 0.001 | 25 635 | 0.391 | |

wGRS indicates weighted genetic risk score; SE, standard error; BMI, body mass index; SBP, systolic blood pressure; HADS, Hospital Anxiety and Depression Scale.
Coefficients are in terms of an average-SNP increase in the allele score per unit/level increase in confounder.
* Associations surpassing multiple-testing corrected p-value threshold of 0.05/64 = 7.81e-04

*Table S15: One-sample Mendelian randomization analysis for risk of incident acute myocardial infarction associated with sleep traits with and without adjustment for potential confounders in UK Biobank and HUNT2.*

| | UK Biobank | | | | HUNT2 | | | |
|---|---|---|---|---|---|---|---|---|
| | MR estimates † | | MR estimates adjusted for potential confounders * | | MR estimates ‡ | | MR estimates adjusted for potential confounders ** | |
| **Sleep trait** | **N (incident cases)** | **Hazard ratio (95% CI)** | **N (incident cases)** | **Hazard ratio (95% CI)** | **N (incident cases)** | **Hazard ratio (95% CI)** | **N (incident cases)** | **Hazard ratio (95% CI)** |
| Insomnia symptoms # | 332 676 (7 813) | 1.18 (1.07, 1.31) | 265 998 (6 098) | 1.04 (0.92, 1.17) | 44 728 (4 488) | 1.23 (1.00, 1.55) | 37 860 (3 425) | 1.13 (0.87, 1.47) |
| 24-hour sleep duration (h) | 332 676 (7 813) | 0.97 (0.75, 1.29) | 265 998 (6 098) | 1.05 (0.76, 1.43) | 44 728 (4 488) | 0.76 (0.31, 1.79) | 37 860 (3 425) | 0.69 (0.29, 1.65) |
| Short sleep # (≤6 h vs. 7-8 h) | 307 135 (7 028) | 1.14 (0.97, 1.32) | 246 136 (5 515) | 1.11 (0.93, 1.32) | 33 243 (3 058) | 0.87 (0.15, 3.24) | 28 776 (2 433) | 0.87 (0.36, 2.07) |
| Long sleep # (≥9 h vs. 7-8 h) | 253 811 (5 762) | 0.83 (0.67, 0.99) | 204 727 (4 526) | 0.87 (0.70, 1.07) | 41 945 (4 209) | 0.53 (0.01, 8.28) | 35 513 (3 209) | 0.59 (0.20, 1.75) |
| Chronotype # (morning preference) | 332 676 (7 813) | 1.06 (0.99, 1.11) | 265 998 (6 098) | 1.06 (0.99, 1.13) | - | - | - | - |

CI indicates confidence interval.

† Derived using unweighted genetic risk score for each sleep trait, with adjustment for age, gender, assessment centre, 40 genetic principal components, and genotyping chip.

‡ Derived using weighted genetic risk score for each sleep trait, with adjustment for age, gender, 20 genetic principal components, and genotyping chip.

* Additionally adjusted for alcohol intake frequency, smoking status, body mass index, physical activity, Townsend deprivation index, education, depression, and chronic illness.

** Additionally adjusted for smoking status, body mass index, serum cholesterol levels, and anxiety.

### Hazard ratio (95% CI) scaled to per doubling in odds of the sleep trait.

*Table S16: Sensitivity analysis for risk of incident acute myocardial infarction associated with sleep traits in UK Biobank.*

| | UK Biobank † | | | | | |
|---|---|---|---|---|---|---|
| **Sleep trait** | **N (incident cases)** | **TSPS HR (95% CI) §** | **IVW HR (95% CI)** | **MR-Egger HR (95% CI)** | **Weighted Median HR (95% CI)** | **Weighted Mode-based HR (95% CI)** |
| Insomnia symptoms # | 332 676 (7 813) | 1.18 (1.07, 1.31) | 1.09 (0.98, 1.20) | 0.77 (0.62, 0.95) ); Intercept 0.007 (0.003, 0.012) | 1.04 (0.91, 1.18) | 1.36 (0.84, 2.19) |
| 24-hour sleep duration (h) | 332 676 (7 813) | 0.97 (0.75, 1.29) | 0.94 (0.70, 1.27) | 0.70 (0.25, 1.97); Intercept 0.005 (-0.012, 0.022) | 0.77 (0.51, 1.16) | 0.38 (0.14, 1.03) |
| Short sleep # (≤6 h vs. 7-8 h) | 307 135 (7 028) | 1.14 (0.97, 1.32) | 1.13 (0.96, 1.33) | 1.09 (0.61, 1.94); Intercept 0.001 (-0.019, 0.022) | 1.24 (0.99, 1.55) | 1.40 (0.88, 2.22) |
| Long sleep # (≥9 h vs. 7-8 h) | 253 811 (5 762) | 0.83 (0.67, 0.99) | 0.83 (0.63, 1.08) | 0.81 (0.40, 1.64); Intercept 0.002 (-0.048, 0.051) | 0.79 (0.61, 1.02) | 0.80 (0.59, 1.09) |
| Chronotype # (morning preference) | 332 676 (7 813) | 1.06 (0.99, 1.11) | 1.06 (0.99, 1.11) | 1.02 (0.88, 1.19); Intercept 0.001 (-0.003, 0.005) | 1.01 (0.94, 1.09) | 0.94 (0.76, 1.17) |

TSPS indicates two-stage predictor substitution; IVW, inverse variance weighted; HR, hazard ratio; and CI, confidence interval.
For IVW, MR-Egger, weighted median and weighted mode-based estimates, the SNP-exposure and the SNP-outcome associations were obtained from the same participants.
† Adjusted for age, gender, assessment centre, 40 genetic principal components, and genotyping chip.
§ Derived using unweighted genetic risk score for each sleep trait
### Hazard ratio (95% CI) scaled to per doubling in odds of the sleep trait

*Table S17: Sensitivity analysis for risk of incident acute myocardial infarction associated with sleep traits in HUNT2.*

| | HUNT2 † | | | | | |
|---|---|---|---|---|---|---|
| **Sleep trait** | **N (incident cases)** | **TSPS HR (95% CI) §** | **IVW HR (95% CI)** | **MR-Egger HR (95% CI)** | **Weighted Median HR (95% CI)** | **Weighted Mode-based HR (95% CI)** |
| Insomnia symptoms # | 44 728 (4 488) | 1.23 (1.00, 1.55) | 1.08 (0.99, 1.17) | 1.16 (1.02, 1.32); Intercept -0.003 (-0.006, 0.001) | 1.12 (0.99, 1.27) | 1.15 (0.87, 1.51) |
| 24-hour sleep duration (h) | 44 728 (4 488) | 0.76 (0.31, 1.79) | 0.71 (0.48, 1.05) | 0.92 (0.47, 1.82); Intercept -0.004 (-0.014, 0.005) | 0.81 (0.45, 1.47) | 0.79 (0.38, 1.65) |
| Short sleep # (≤6 h vs. 7-8 h) | 33 243 (3 058) | 0.87 (0.15, 3.24) | 1.06 (0.88, 1.29) | 1.07 (0.75, 1.52); Intercept -0.001 (-0.015, 0.014) | 1.09 (0.80, 1.50) | 1.07 (0.81, 1.42) |
| Long sleep # (≥9 h vs. 7-8 h) | 41 945 (4 209) | 0.53 (0.01, 8.28) | 1.07 (0.61, 1.90) | 0.95 (0.38, 2.39); Intercept 0.006 (-0.030, 0.042) | 0.94 (0.50, 1.75) | 1.02 (0.63, 1.65) |

TSPS indicates two-stage predictor substitution; IVW, inverse variance weighted; HR, hazard ratio; and CI, confidence interval.
For IVW, MR-Egger, weighted median and weighted mode-based estimates, the SNP-exposure and the SNP-outcome associations were obtained from the same participants.
† Adjusted for age, gender, 20 genetic principal components, and genotyping chip.
§ Derived using weighted genetic risk score for each sleep trait.
### Hazard ratio (95% CI) scaled to per doubling in odds of the sleep trait.

*Table S18: One-sample Mendelian randomization Cox regression analysis for risk of incident acute myocardial infarction associated with insomnia symptoms using instruments from Lane et al., 2019 [16] in UK Biobank and HUNT2.*

| | **N (incident cases)** | **TSPS HR (95% CI)** § | **IVW HR (95% CI)** | **MR-Egger HR (95% CI)** | **Weighted Median HR (95% CI)** | **Weighted Mode-based HR (95% CI)** |
|---|---|---|---|---|---|---|
| **UK Biobank** † | 332 676 (7 813) | 1.25 (1.09, 1.45) | 1.12 (0.96, 1.31) | 0.69 (0.50, 0.96); Intercept 0.013 (0.005, 0.022) | 1.18 (0.96, 1.44) | 1.15 (0.83, 1.60) |
| **HUNT2** ‡ | 44 728 (4 488) | 1.39 (0.97, 2.41) | 1.13 (0.98, 1.29) | 0.97 (0.78, 1.19); Intercept 0.006 (0.001, 0.012) | 1.11 (0.90, 1.37) | 1.14 (0.92, 1.41) |

TSPS indicates two-stage predictor substitution; IVW, inverse variance weighted; HR, hazard ratio; and CI, confidence interval.
Hazard ratio (95% CI) scaled to per doubling in odds of the sleep trait.
For IVW, MR-Egger, weighted median and weighted mode-based estimates, the SNP-exposure and the SNP-outcome associations were obtained from the same participants.
§ Derived using unweighted genetic risk score for insomnia symptoms in UK Biobank and weighted genetic risk score for insomnia symptoms in HUNT2.
† Adjusted for age, gender, assessment centre, 40 genetic principal components, and genotyping chip.
‡ Adjusted for age, gender, 20 genetic principal components, and genotyping chip.

*Table S19: Sensitivity analysis for risk of incident acute myocardial infarction associated with insomnia symptoms and chronotype in UK Biobank using genetic variants genome-wide significant in 23andMe.*

| | UK Biobank † | | | | | | |
|---|---|---|---|---|---|---|---|
| **Sleep trait** | **N (incident cases)** | **TSPS HR (95% CI) §** | **TSPS HR (95% CI) *** | **IVW HR (95% CI)** | **MR-Egger HR (95% CI)** | **Weighted Median HR (95% CI)** | **Weighted Mode-based HR (95% CI)** |
| Insomnia symptoms # | 332 676 (7 813) | 1.25 (1.08, 1.45) | 1.33 (1.13, 1.56) | 1.16 (0.99, 1.35) | 0.78 (0.59, 1.02); Intercept 0.008 (0.003, 0.013) | 1.10 (0.89, 1.35) | 1.01 (0.61, 1.65) |
| Chronotype # (morning preference) | 332 676 (7 813) | 1.02 (0.94, 1.11) | 0.99 (0.91, 1.09) | 1.02 (0.92, 1.13) | 1.14 (0.91, 1.43); Intercept -0.004 (-0.013, 0.004) | 0.95 (0.84, 1.08) | 0.92 (0.71, 1.18) |

TSPS indicates two-stage predictor substitution; IVW, inverse variance weighted; HR, hazard ratio; and CI, confidence interval.
For IVW, MR-Egger, weighted median and weighted mode-based estimates, the SNP-exposure and the SNP-outcome associations were obtained from the same participants.
† Adjusted for age, gender, assessment centre, 40 genetic principal components, and genotyping chip.
§ Derived using weighted genetic risk score for each sleep trait.
* Derived using unweighted genetic risk score for each sleep trait.
### Hazard ratio (95% CI) scaled to per doubling in odds of the sleep trait

*Table S20: Sensitivity analysis for risk of incident acute myocardial infarction associated with insomnia symptoms in HUNT2 using genetic variants genome-wide significant in 23andMe.*

| | HUNT2 † | | | | | | |
|---|---|---|---|---|---|---|---|
| **Sleep trait** | **N (incident cases)** | **TSPS HR (95% CI) §** | **TSPS HR (95% CI) *** | **IVW HR (95% CI)** | **MR-Egger HR (95% CI)** | **Weighted Median HR (95% CI)** | **Weighted Mode-based HR (95% CI)** |
| Insomnia symptoms # | 44 728 (4 488) | 1.15 (0.90, 1.46) | 1.10 (0.85, 1.42) | 1.11 (1.00, 1.25) | 1.23 (1.03, 1.47); Intercept: -0.004 (-0.009, 0.001) | 1.12 (0.95, 1.32) | 1.18 (0.93, 1.51) |

TSPS indicates two-stage predictor substitution; IVW, inverse variance weighted; HR, hazard ratio; and CI, confidence interval.
For IVW, MR-Egger, weighted median and weighted mode-based estimates, the SNP-exposure and the SNP-outcome associations were obtained from the same participants.
† Adjusted for age, gender, 20 genetic principal components, and genotyping chip.
§ Derived using weighted genetic risk score for insomnia symptoms.
* Derived using unweighted genetic risk score for insomnia symptoms.
### Hazard ratio (95% CI) scaled to per doubling in odds of the sleep trait.

*Table S21: List of medications used to define the sleep medication covariate in UK Biobank.*

| **Sleep medication** | **Treatment/medication code** (UK Biobank field ID: 20003) |
|---|---|
| Oxazepam | 1140863442 |
| Meprobamate | 1140863378 |
| Medazepam | 1140863372 |
| Bromazepam | 1140863318 |
| Lorazepam | 1140863302 |
| Clobazam | 1140863268 |
| Chlormezanone | 1140863262, 1140868274 |
| Temazepam | 1140863202 |
| Nitrazepam | 1140863182, 1140863104 |
| Lormetazepam | 1140863176 |
| Diazepam | 1140863152, 1141157496 |
| Zopiclone | 1140863144 |
| Triclofos sodium | 1140863140 |
| Methyprylone | 1140856040 |
| Prazepam | 1140855944 |
| Triazolam | 1140855914 |
| Ketazolam | 1140855860 |
| Dichloralphenazone | 1140855824 |
| Clomethiazole | 1140909798 |
| Zaleplon | 1141171404 |
| Butobarbital | 1141180444 |
| Clonazepam | 1140872150 |
| Flurazepam | 1140863110 |
| Loprazolam | 1140863120 |
| Alprazolam | 1140863308 |
| Butobarbitone | 1140882090 |
